# Mast cell score associates with wide-spread mast cell symptoms and comorbidities in patients with hEDS and HSD

**DOI:** 10.64898/2026.05.31.26354552

**Authors:** Frances C. Wilson, Derek J. Zangerle, Lincoln E. Rozen, Jessica J. Fliess, Ashley A. Darakjian, Keith A. Sacco, Charwan Hamilton, Max W. Strandes, Alayna M. Puls, Cameron J. Hartmoyer, Sophia N. Witola Reyes, Stacey M. Menton, Daniel V. Dudenkov, Alexei Gonzalez-Estrada, Sarah C. Solomon, Ina Stephens, Benjamin W.E. Wang, Paldeep S. Atwal, Chrisandra L. Shufelt, Rachel M. Botella, Ashley M. Zeman, Dacre R.T. Knight, Shilpa N. Gajarawala, Katelyn A. Bruno, DeLisa Fairweather

**Affiliations:** Department of Cardiovascular Medicine, Mayo Clinic, Jacksonville, Florida, USA; Division of General Internal Medicine, Department of Internal Medicine, Mayo Clinic, Jacksonville, Florida, USA; Division of Cardiovascular Medicine, University of Florida, Gainesville, Florida, USA; Department of Allergy, Pulmonary and Sleep Medicine, Mayo Clinic, Jacksonville, Florida, USA; Department of Otolaryngology-Head and Neck Surgery, Mayo Clinic, Jacksonville, Florida, USA; Division of Allergy, Asthma and Clinical Immunology, Department of Medicine, Mayo Clinic, Scottsdale, Arizona, USA; Department of Pediatrics, University of Virginia, Charlottesville, Virginia, USA; Division of Rheumatology, Department of Internal Medicine, Mayo Clinic, Jacksonville, Florida, USA; Atwal Clinic, West Palm Beach, Florida, USA; Department of Medicine, University of Virginia, Charlottesville, Virginia, USA; Department of Immunology, Mayo Clinic, Jacksonville, Florida, USA; Center for Clinical and Translational Science, Mayo Clinic, Rochester, Minnesota, USA

**Author notes:** **Correspondence: DeLisa Fairweather, PhD,**. **Co-First authorship:** FCW, DJZ, LER contributed equally to this work and share first authorship. **Equal contribution and senior authorship:** DRTK, SNG, KAB and DF contributed equally to this work and share senior authorship.

**Keywords:** hypermobile Ehlers-Danlos syndrome, hypermobility spectrum disorders, mast cells, mast cell activation syndrome, mast cell activation disorders, abuse, adverse childhood experiences

## Abstract

**Background:** Wide-spread mast cell (MC)-associated symptoms and MC activation syndrome (MCAS) are often reported in patients with hypermobile Ehlers-Danlos syndrome (hEDS) and hypermobility spectrum disorders (HSD). The goal of this study was to develop a novel MC score based on 11 self-reported MC-related conditions with clinical and research utility to better understand MC symptoms in hEDS and HSD patients.

**Methods:** From November 1, 2019, to June 13, 2025, patients (*n*=2,141) filled out an Intake Questionnaire at the Mayo Clinic Florida EDS Clinic that included 11 self-reported questions related to categories of MC-related conditions for a MC score ranging from 0/11 to 11/11. Based on the MC score distribution in hEDS and HSD patients, a MC score of 0-1 was considered a low MC score and ≥5 was considered a high MC score. Symptoms/comorbidities were compared between patients with high vs. low MC scores.

**Results:** From the 2,141 hEDS/HSD patients, 535 (25.0%) had a MC score ≥5 (Hi MC). MCAS-specific symptoms such as nausea and vomiting were reported more often in hEDS/HSD patients with a high vs. low MC score (*p*<0.0001). Random clinical blood tryptase and urinary MC markers were not elevated in patients with high MC scores (*n*=50/group), although high MC scores were found to significantly reduce urinary creatinine levels indicating that the protein used to normalize data was affected by MC activity. In contrast, random blood IgE, tryptase and major basic protein (MBP) by ELISA were increased in patients with high MC scores (e.g., IgE hEDS *p*=0.0004, HSD *p*=0.003). Of note, the percentage of patients reporting abuse or post-traumatic stress disorder was nearly doubled in patients with high vs. low MC scores (Abuse and PTSD: hEDS *p* < 0.0001; HSD *p* < 0.0001). Overall, 109/135 (80.7%) in hEDS and 129/135 (95.6%) in HSD reported more symptoms/comorbidities if they had a high MC score.

**Conclusions:** We found that hEDS/HSD patients with high MC scores self-reported more widespread symptoms/comorbidities and higher MC-related blood markers than patients with low MC scores indicating the utility of this tool to evaluate the level of widespread MC activity in hEDS, HSD and other patients.

## 1 Introduction

Hypermobile Ehlers-Danlos syndrome (hEDS) and hypermobility spectrum disorders (HSD) are heritable musculoskeletal disorders with widespread distribution of fragile connective tissue in the skin, joints, ligaments, and internal organs.^1^ Based on survey data, 255 million people worldwide (3%) are estimated to have hEDS or HSD.^2^ Symptoms and comorbidities are often multiorgan, multifaceted, and potentially debilitating including greater joint pain, musculoskeletal pain, and fatigue than the general population.^3^ In classical forms of EDS gene variants have been identified in collagen or other extracellular matrix (ECM) proteins.^4^ In contrast, gene variants unique to hEDS or HSD are unknown except for several recent reports including for variants in patients with hEDS in the kallikrein gene family.^5,6^ Kallikrein genes encode serine proteases which are found in a number of cells/tissues including mast cells (MCs).^5^ MCs are critical for mounting allergic responses, ECM remodeling and drive many tissue-related inflammatory conditions.^7,8^ So, genetic variants that alter MC function may explain the similarity in systemic and connective tissue issues as well as widespread symptoms in many different tissues in patients with hEDS/HSD.

Many clinicians have observed heightened MC and widespread activity in patients with hEDS and HSD based primarily on symptoms.^9–12^ For a diagnosis of MC activation syndrome (MCAS) the following criteria must be met: 1. episodic symptoms consistent with MC degranulation involving at least 2 organ systems (i.e., cutaneous, GI, cardiovascular, or respiratory), 2. event-related increase in serum tryptase above the individual’s baseline tryptase to 20% + 2ng/mL, and 3. response of symptoms to therapy with MC-stabilizing agents, drugs directed against MC mediator production, or drugs blocking mediator release or effects of MC-derived mediators, and 4. the symptoms were not better explained by another condition.^13–15^ MC activation disorders (MCAD) are defined as patients with MCAS, mastocytosis (cKit mutation) and/or a tryptase mutation (hereditary alpha tryptasemia/HAT).^14^ One of the current debates in the EDS literature is whether MCAS is rare or a relatively common condition.^10,15–22^ The current diagnostic criteria for MCAS has been referred to as ‘consensus 1’, while a group of investigators and physicians have developed a ‘consensus 2’ definition for MCAS that is inclusive of the many MC-related symptoms throughout the body without the requirement for elevated tryptase or reversal of symptoms with medication.^17^ Wang et al. reported that of 195 patients seen for autonomic dysfunction that 8.2% were diagnosed with EDS, postural orthostatic tachycardia syndrome (POTS) and MCAS.^11^ Within the group diagnosed with POTS and EDS, 31% were diagnosed with MCAS compared to 2% in the non-POTS/EDS group.^11^ Of 974 patients with suspected MCAD, Brock et al. found that 46% had a diagnosis of MCAD and 10% had hEDS/HSD and MCAD.^10^ However, studies often do not clarify whether they were using ‘consensus 1’ or ‘consensus 2’ for their assessment of MCAS.^22^ A study by Yao et al. examined children and adults with a diagnosis of POTS for the percentage of hEDS/HSD and MCAS using their own modified diagnostic criteria for hEDS.^23^ From 100 patients with POTS, MCAS ranged from 2% using ‘consensus 1’ criteria, 37% using ‘consensus 2’ criteria, and 87% based on the presence of MC-related symptoms.^23^ The true prevalence of MCAS and/or MCAD in the hEDS/HSD population is unknown and prevalence estimates are complicated by application of different diagnostic criteria.

In this study we developed a novel MC score with the aim of providing a clinical and research tool that reflects widespread MC issues in patients rather than an acute/anaphylactic response to an allergic stimulus, as is required for a diagnosis of MCAS using ‘consensus 1’. The goal was to develop a tool that would quantitate the level of widespread MC activation in patients that could be used by physicians and researchers to gauge the severity of disease and response to therapies. The MC score is intended to be used in addition to the MCAS (consensus 1) criteria. The MC score is based on patients self-reporting whether they experience 11 MC-related conditions including environmental allergies, asthma (past and/or present), atopy, food allergies, venom/stinging insect allergies, rhinitis, eczema, drug/chemical allergies/intolerance, MCAS, MC hyperactivity, and/or a tryptase mutation. Each ‘yes’ answer is given a value of 1 so that patients have a MC score ranging from 0/11 to 11/11, similar in principal to the adverse childhood experiences (ACE) score.^24^ we also examined the relationship of the MC score to the ACE score. We *hypothesized* that MCAS using criteria 1 would represent a small subset of hEDS/HSD patients with a high MC score. We found that around 25% of patients with hEDS/HSD in this study reported widespread MC activation issues based on a MC score ≥5 (a high MC score), which was associated with worse symptoms/comorbidities for almost all widespread symptoms/comorbidities that we examined and elevated blood MC markers. We also found that the percentage of patients reporting abuse or post-traumatic stress disorder (PTSD) was nearly doubled in patients with high MC scores which was reflected in higher ACE scores.

## 2. Methods

### 2.1 Ethics statement

The Institutional Review Board (IRB) of Mayo Clinic approved the retrospective analysis (IRB# 19-010260) of demographic and clinical data from medical records for this study and waived informed consent for all patients. Analysis of patient samples for research purposes was approved using IRB# 19-011260 after obtaining consent. The research conformed to the principles outlined in the Declaration of Helsinki.

### 2.2 Diagnosis of hEDS and HSD patients

Patients were seen at the Mayo Clinic Florida EDS Clinic by self-referral or referrals from inside or outside Mayo Clinic. Patients were diagnosed with hEDS according to the 2017 diagnostic criteria,^4^ as previously.^25–28^ Briefly, the diagnostic criteria for hEDS includes identification of generalized joint hypermobility (GJH) of specific joints using the Beighton Scale (past puberty ≥5/9 and over 50 years of age ≥4/9), evidence of a systemic connective tissue disorder, family history and/or musculoskeletal complications, and several exclusions.^4^ HSD is diagnosed in patients that do not meet the diagnostic criteria for hEDS but have a positive Beighton Score (≥5/9 and ≥4/9 for older patients) and evidence that the joint hypermobility is causing problems and it is not just an asymptomatic feature (feature C of the 2^nd^ EDS criterion).^4,29^ This study did not include patients with L-HSD, H-HSD, or other forms of HSD or other connective tissue disorders.^28^

### 2.3 EDS Clinic data collection

Patients received a clinical Intake Questionnaire as standard of care prior to their first appointment at the Mayo Clinic Florida EDS Clinic. The Intake Questionnaire was originally designed by Drs. Bruno and Fairweather based on patient symptoms.^25,30^ Self-reported data on 135 symptoms/comorbidities were obtained from the Intake Questionnaire of adult patients >18 years of age from November 1, 2019, to June 13, 2025, (*n*=2,141) with a valid mast cell score. The Intake Questionnaire categorized questions by organ or system such as muscles, joints, allergy, neurological symptoms or gastrointestinal (GI) symptoms, as previously.^25,27,28^ Thus, data were organized in the manuscript according to organ or system categories.

### 2.4 Mast cell score

The mast cell (MC) score was developed by Drs. Fairweather and Bruno and is based on a range of known MC responses (i.e., allergy, atopy, asthma). A higher MC score indicates more widespread MC activity. To obtain the MC score patients were asked a series of 11 questions related to categories of MC activity that included: 1. Do you have any environmental allergies? 2. Do you have a venom (i.e., snake bite) or stinging insect (i.e., bee) allergy? 3. Do you have any food allergies? 4. Do you have any drug/chemical allergies? 5. Do you have allergic rhinitis, otherwise known as hay fever? 6. Do you have atopic dermatitis (eczema)? 7. Do you have atopy (a hypersensitivity reaction that occurs in any area of the body that is not in contact with the allergen; often occurs on skin)? 8. Do you currently have or have ever had asthma? Have you been told that you have a hyper immune response like mast cell activation including: 9. Mast cell activation syndrome (MCAS)? 10. Overactive mast cells? 11. Tryptase mutation? A minimum of 10 of the 11 questions were required to be answered for a valid score. Each ‘yes’ answer was given a value of ‘1’ so that the MC score ranged from 0/11 to 11/11 and is shown as an average ±SD comparing groups. Self-reported MC data were not validated by another method such as checking the medical record for a diagnosis code. Additionally, based on the distribution of MC scores by diagnosis (**Fig.1a**), MC scores of 0-1 were chosen to represent low MC scores (Lo MC) and MC scores of ≥5 were chosen to represent high MC scores (Hi MC). After exclusion of patients with MC scores ranging from 2-4, we examined a total of 1,050 patients (**Fig.1b**).

### 2.5 Blood and urine MC markers

Blood and urine samples were collected from patients to examine various MC markers for clinical purposes on the day of their clinic appointment and so were ‘random’ samples, because they were not collected in response to an allergic reaction. Random blood samples were also collected on the clinic appointment day for those who had consented to research. Clinical samples were sent to Mayo Clinic Clinical Laboratories for assessment of tryptase and urine MC markers including N-methylhistamine, 2,3-dinor-11β-prostaglandin (PG) F2α, and leukotriene E4 all normalized to creatinine (Cr). The Fairweather lab performed ELISAs on sera from blood samples acquired prospectively using the following human kits: tryptase (Innovative Research, Inc., Novi, MI, Cat# IHUTPSKT, detection limit <15 pg/mL), total IgE (Thermo Fischer Scientific, Waltham, MA, Cat# BMS2097, detection limit 0.5 ng/mL), and major basic protein (MBP) (MyBioSource, San Diego, CA, Cat# MBS2019468, detection limit <1.25 ng/mL).

### 2.6 Adverse childhood experiences score

The Adverse Childhood Experiences (ACE) score is a cumulative measure of traumatic or stressful experiences occurring before age 18.^31^ As a part of the EDS Clinic Intake Questionnaire, we asked the following 10 ACE questions: 1. Did a parent or other adult in the household often or very often: a) Swear at you, insult you, put you down, or humiliate you? or b) Act in a way that made you afraid that you might be physically hurt? 2. Did a parent or other adult in the household often or very often: a) Push, grab, slap, or throw something at you? or b) Ever hit you so hard that you had marks or were injured? 3. Did an adult or person at least 5 years older than you ever: a) Touch or fondle you or have you touch their body in a sexual way? or b) Attempt or actually have oral, anal, or vaginal intercourse with you? 4. Did you often or very often feel that: a) No one in your family loved you or thought you were important or special? or b) Your family didn’t look out for each other, feel close to each other, or support each other? 5. Did you often or very often feel that: a) You didn’t have enough to eat, had to wear dirty clothes, and had no one to protect you? or b) Your parents were too drunk or high to take care of you or take you to the doctor if you needed it? 6. Were your parents ever separated or divorced? 7. Was your parent/caregiver: a) Often or very often pushed, grabbed, slapped or had something thrown at him/her? or b) Sometimes, often, or very often kicked, bitten, hit with a fist, or hit with something hard? or c) Ever repeatedly hit over at least a few minutes or threatened with a gun or knife? 8. Did you live with anyone who was a problem drinker or alcoholic, or who used street drugs? 9. Was a household member depressed or mentally ill, or did a household member attempt suicide? 10. Did a household member go to prison? Each category with an answer of ‘yes’ adds 1 point to the total ACE score for a range of 0/10 to 10/10. Higher ACE scores indicate that an individual experienced a larger number of childhood adversities, which has been associated with an increased risk for later health and social problems.^24,32,33^

### 2.7 Statistical analysis

Missing data were removed from the assessment and the number of patients per group shown for individual questions or sets of questions within tables. Student’s *t* test (for parametric data) or Mann-Whitney rank test (for nonparametric data) was used to compare continuous variables between two groups. Age and BMI were summarized as median and interquartile range. Categorical variables were summarized in tables as number and percentage of subjects. Although Fisher’s exact test has been historically limited to small sample sizes, the current Prism statistical package overcomes this limitation so that Fisher’s exact test is more accurate than a Chi squared test for large numbers. Thus, we used Fisher’s exact test to compare categorical variables between two groups and to determine odds ratios with 95% confidence intervals. The specific tests that were used are listed in the legends of the tables and figures. All the tests were two-tailed and *p* values <0.05 were considered statistically significant. We examined 135 symptoms/comorbidities in this study. This was an exploratory study, and so uncorrected p values are shown in the tables and figures. Correction for multiple comparisons indicates that only *p* values <0.0004 were significant. All statistical analyses were performed using Microsoft Excel and GraphPad Prism (Version 10.6.0).

## Results

### 3.1 Patient demographics

From the 2,141 hEDS/HSD patients who were assessed in the study, we found that 515 (24.1%) had a MC score of 0-1 (Lo MC), 1,091 (51.0%) had a MC score of 2-4 (removed from analysis), and 535 (25.0%) had a MC score ≥5 (Hi MC) (**Fig1ab**). From the total population, 500 (23.4%) had a diagnosis of hEDS and 1,641 (76.7%) had a diagnosis of HSD. From the total population, 144 (6.7%) had a diagnosis of hEDS and a high MC score while 391 (18.3%) had a diagnosis of HSD and a high MC score (**Table 1**).

**Table 1.** Demographics of patients diagnosed with hEDS or HSD with low (Lo) vs. high (Hi) MC scores (*n* = 1,044)

| Condition | hEDS<sup>a</sup><br>Lo MC<br>$n$ (%) | hEDS<br>Hi MC<br>$n$ (%) | $P$ value<sup>b</sup> | HSD<br>Lo MC<br>$n$ (%) | HSD<br>Hi MC<br>$n$ (%) | $P$<br>value<sup>b</sup> |
| --- | --- | --- | --- | --- | --- | --- |
| **Age** | $n = 114$ | $n = 143$ | **<0.0001** | $n = 398$ | $n = 388$ | **<0.0001** |
| Median (interquartile range) | 26.1<br>(21.3-38.6) | 36.5<br>(29.1-45.7) |  | 27.3<br>(21.5-37.0) | 36.7<br>(26.2-45.6) |  |
| **Sex** | $n = 115$ | $n = 141$ | | $n = 400$ | $n = 388$ | |
| Female | 104 (90.4) | 126 (89.4) | 0.84 | 366 (91.5) | 355 (91.5) | 0.99 |
| Male | 9 (7.8) | 11 (7.8) | 0.99 | 29 (7.3) | 24 (6.2) | 0.57 |
| Non-binary | 2 (1.7) | 3 (2.1) | 0.99 | 5 (1.3) | 8 (2.1) | 0.41 |
| Other | 0 (0.0) | 1 (0.7) | 0.99 | 0 (0.0) | 1 (0.3) | 0.49 |
| Unknown | 0 (0.0) | 0 (0.0) | 0.99 | 0 (0.0) | 0 (0.0) | 0.99 |
| **Race** | $n = 115$ | $n = 144$ | | $n = 400$ | $n = 390$ | |
| American Indian/<br>Alaskan Native | 2 (1.7) | 2 (1.4) | 0.99 | 2 (0.5) | 10 (2.6) | **0.020** |
| Asian | 2 (1.7) | 3 (2.1) | 0.99 | 8 (2.0) | 5 (1.3) | 0.58 |
| Black or African<br>American | 2 (1.7) | 4 (2.8) | 0.69 | 6 (1.5) | 16 (4.1) | **0.030** |
| Native Hawaii/<br>Pacific Islander | 0 (0.0) | 0 (0.0) | 0.99 | 1 (0.3) | 0 (0.0) | 0.99 |
| White | 106 (92.2) | 128 (88.9) | 0.41 | 379 (94.8) | 362 (92.8) | 0.30 |
| Other | 3 (2.6) | 8 (5.6) | 0.36 | 8 (2.0) | 14 (3.6) | 0.19 |
| Unknown/ not disclosed | 2 (1.7) | 3 (2.1) | 0.99 | 8 (2.0) | 8 (2.1) | 0.99 |
| **Ethnicity** | 115 | 144 |  | 400 | 390 |  |
| Hispanic or Latino | 13 (11.3) | 9 (6.3) | 0.18 | 35 (8.8) | 35 (9.0) | 0.99 |
| Not Hispanic or Latino | 100 (87.0) | 129 (89.6) | 0.56 | 350 (87.5) | 344 (88.2) | 0.83 |
| Unknown/ not disclosed | 2 (1.7) | 6 (4.2) | 0.31 | 15 (3.8) | 11 (2.8) | 0.55 |
| **Highest level of education** | *n* = 115 | *n* = 135 |  | *n* = 400 | *n* = 390 |  |
| Some high school | 9 (7.8) | 1 (0.7) | **0.007** | 32 (8.0) | 10 (2.6) | **0.0007** |
| High school/ GED | 18 (15.7) | 10 (7.4) | **0.045** | 53 (13.3) | 25 (6.4) | **0.001** |
| Some college | 32 (27.8) | 25 (18.5) | 0.09 | 82 (20.5) | 78 (20.0) | 0.93 |
| Trade school | 2 (1.7) | 0 (0.0) | 0.21 | 15 (3.8) | 26 (6.7) | 0.08 |
| Associate's degree | 11 (9.6) | 23 (17.0) | 0.09 | 37 (9.3) | 39 (10.0) | 0.81 |
| Bachelor's degree | 31 (27.0) | 37 (27.4) | 0.99 | 113 (28.3) | 114 (29.2) | 0.81 |
| Master's degree | 7 (6.1) | 26 (19.3) | **0.002** | 47 (11.8) | 72 (18.5) | **0.01** |
| Professional/ Doctorate | 5 (4.3) | 13 (9.6) | 0.14 | 17 (4.3) | 24 (6.2) | 0.26 |
| Not disclosed | 0 (0.0) | 0 (0.0) | 0.99 | 4 (1.0) | 2 (0.5) | 0.69 |

<sup>a</sup> All data shown as *n* (%). <sup>b</sup> P values obtained from Fisher's test for categorical data and Mann-Whitney test for numeric data.

The median and interquartile age range of hEDS patients with a low MC score was 26.1 (21.3-38.6) vs. with a high MC score was 36.5 (29.1-45.7) (*p* < 0.0001) (**Table 1**). Similarly, the age range of HSD patients with a low MC score was 27.3 (21.5-37.0) vs. with a high MC score was 36.7 (26.2-45.6) (*p* < 0.0001) (**Table 1**). Thus, patients with high MC scores were nearly a decade older than those with low MC scores.

Over 90% in each group self-reported as females (**Table 1**). Most patients in each category self-reported as non-Hispanic White; however, more HSD patients with Hi MC activity (3.8%) self-reported as Black/African American than HSD patients with Lo MC activity (1.5%) (*p* = 0.042) (**Table 1**). Additionally, patients with Hi MC activity generally reported receiving higher education degrees like Masters and PhDs compared to patients with Lo MC activity (**Table 1**).

The median and interquartile BMI range of hEDS patients with a Lo MC score was 22.4 (20.3-26.1) vs. a Hi MC score was 24.6 (21.3-29.1) (*p* = 0.024) while the BMI of HSD patients with a Lo MC score was 24.4 (21.3-29.7) vs. those with a Hi MC score was 27.5 (23.3-33.0) (*p* < 0.0001) (**Supplemental Table 1**). Thus, patients with high MC scores had a higher BMI, but none of the groups were obese.

hEDS patients with Hi MC activity smoked more cigarettes/day (10-20 cigarettes Lo MC 7.6% vs. Hi MC 25.9%, *p* = 0.018), consumed more alcohol (1/week Lo MC 54.4% vs. Hi MC 70.9%, *p* = 0.026), and were illicit drug users in the past (Lo MC 1.4% vs. Hi MC10.1%, *p* = 0.0006) compared to hEDS patients with Lo MC activity (**Supplemental Table 1**). Similarly, HSD patients with Hi MC activity had a past smoking history (Lo MC 10.8% vs. 18.4%, *p* = 0.001), smoked more cigarettes/day (6-10 cigarettes Lo MC 3.3% vs. Hi MC 11.0%, *p* = 0.041), and had secondary smoke exposure (Lo MC 35.5% vs. Hi MC 47.3%, *p* = 0.0003) compared to HSD patients with Lo MC activity, but they had less current illicit drug use (Lo MC 6.6% vs. Hi MC 3.4%, *p* = 0.033) (**Supplemental Table 1**). Thus, hEDS/HSD patients with high MC scores had more exposure to cigarette smoke.

### 3.2 Mast cell score

Figure 1a shows the distribution of MC scores among patients with hEDS or HSD, which were similar between the two diagnoses. The mean MC score ±SD for hEDS patients was 3.01 ±1.95 and for HSD was 3.04 ±1.94 (**Fig1ab**). Patients with MC scores of 0-1 (24.1%) were selected to represent Lo MC responses and patients with MC scores ≥5 (25.0%) were selected to represent patients with high MC scores (**Fig1b**), which produced similar percentages of patients for the analysis by group. Patients with scores ranging from 2-4 were removed from the analysis (**Fig1b**).

**Figure 1.**
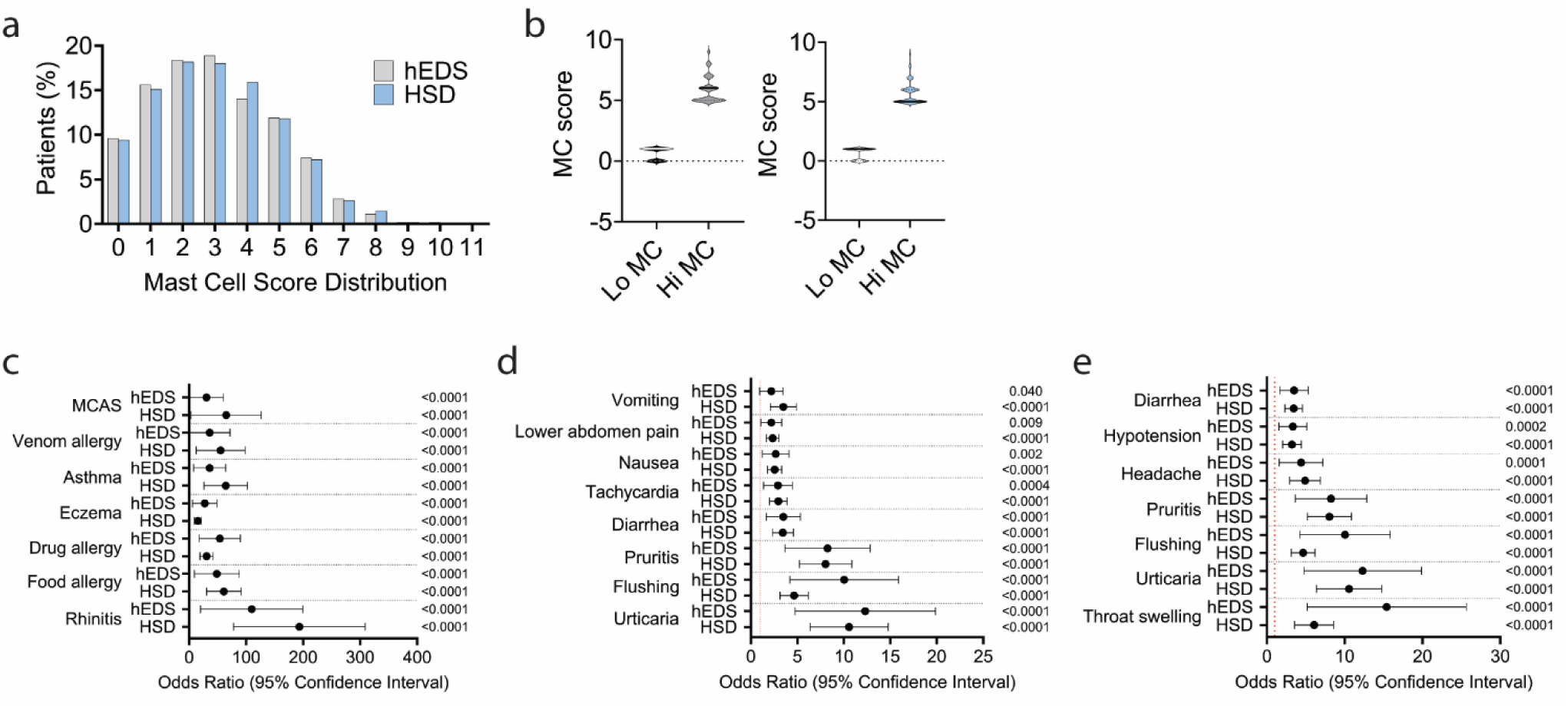
Patients with a high MC score self-report more MC-related symptoms. a) Distribution of MC score for 11 self-reported MC-associated conditions in patients with hEDS (grey) and HSD (blue). b) Distribution of patients with a low (0-1, Lo MC) or high (5-11, High MC) score. Patients with a MC score of 2-4 were removed from the study. c) Odds ratios (OR) and 95% confidence interval (CI) comparing hEDS or HSD patients with Lo vs. High MC scores for self-reported conditions included in the MC score (several not shown due to high CI). d) OR and CI for MC-related symptoms/conditions associated with MCAS according to Akin et al.^13^ e) OR and CI for MC-related symptoms/conditions associated with MCAS according to Valent et al.^14^ c-e) P values obtained from Fisher’s exact test. Only P values less than 0.006 were significant after correcting for multiple comparisons.

Table 2 lists the number and percentage of hEDS and HSD patients with self-reported MC-related symptoms/comorbidities that were used to create the MC score. Patients with a Hi MC score reported more MC score-related symptoms than patients with a Lo MC score (**Table 2**, **Fig1c**). The top three MC-related conditions reported by hEDS/HSD patients were environmental allergies (hEDS OR 439.3, CI 107.1-1859, *p* < 0.0001; HSD OR 128.3, CI 68.86-244.9, *p* < 0.0001), drug allergies (hEDS OR 46.34, CI 21.93-92.94, *p* < 0.0001; HSD OR 29.05, CI 19.77-42.92, *p* < 0.0001) and rhinitis (hEDS OR 87.24, CI 33.59-208.5, *p* < 0.0001) (**Table 2**, **Supplemental Table 2**, **Fig1c**).

**Table 2.** Eleven self-reported MC-related symptoms that were used to obtain MC scores in patients diagnosed with hEDS or HSD with low (Lo) vs. high (Hi) MC scores (*n* = 1,050)

| Condition | hEDS<sup>a</sup><br>Lo MC<br>(*n* = 115) | hEDS<br>Hi MC<br>(*n* = 144) | *P* value<sup>b</sup> | HSD<br>Lo MC<br>(*n* = 400) | HSD<sup>c</sup><br>Hi MC<br>(*n* = 391) | *P* value<sup>b</sup> |
| --- | --- | --- | --- | --- | --- | --- |
| Environmental allergies | 16 (13.9) | 142 (98.6) | **<0.0001<sup>d</sup>** | 79 (19.8) | 379 (96.9) | **<0.0001** |
| Atopy | 1/40 (2.5) | 55/59<br>(93.2) | **<0.0001** | 4/52 (7.7) | 63/70<br>(90.0) | **<0.0001** |
| Drug allergies | 18 (15.7) | 129 (89.6) | **<0.0001** | 72 (18.0) | 338 (86.4) | **<0.0001** |
| Rhinitis (i.e., hayfever) | 5 (4.3) | 115 (79.9) | **<0.0001** | 12 (3.0) | 331 (84.7) | **<0.0001** |
| Food allergies | 5 (4.3) | 92 (63.9) | **<0.0001** | 19 (4.8) | 287 (73.4) | **<0.0001** |
| Asthma | 6 (5.6) | 80 (63.5) | **<0.0001** | 12 (3.2) | 226 (65.3) | **<0.0001** |
| Eczema | 7 (6.1) | 84 (58.3) | **<0.0001** | 31 (7.8) | 215 (55.0) | **<0.0001** |
| Venom allergies | 3 (2.6) | 57 (39.6) | **<0.0001** | 5 (1.3) | 141 (36.1) | **<0.0001** |
| MCAS | 3 (2.6) | 51 (35.4) | **<0.0001** | 3 (0.8) | 112 (28.6) | **<0.0001** |
| Overactive mast cells | 0 (0.0) | 17 (11.8) | **<0.0001** | 1 (0.3) | 53 (13.6) | **<0.0001** |
| Tryptase mutation | 0 (0.0) | 2 (1.4) | 0.50 | 0 (0.0) | 6 (1.5) | **0.014** |
| Other MC disorder | 5 (4.3) | 14 (9.7) | 0.15 | 10 (2.5) | 38 (9.7) | **<0.0001** |
| No MC issues | 40 (34.8) | 14 (9.7) | **<0.0001** | 169 (42.3) | 22 (4.3) | **<0.0001** |
| Unknown | 67 (58.3) | 59 (41.0) | **0.006** | 213 (53.3) | 194 (49.6) | 0.31 |

<sup>a</sup> All data shown as *n* (%). <sup>b</sup> P values obtained from Fisher's exact test for categorical data. <sup>c</sup> Order of conditions based on highest to lowest percentage in HSD Hi MC group. <sup>d</sup> Bold indicates significant value *p* < 0.05; values higher than *p* < 0.0004 were not significant after Bonferroni correction.

### 3.3 MCAS-related symptoms and comorbidities

Table 3 lists MC-related symptoms and comorbidities that have been associated with MCAS by Akin et al. and Valent et al. as part of the diagnostic criteria for MCAS (consensus 1).^13,14^ We found that more patients with a Hi MC score self-reported these symptoms than patients with a Lo MC score for both diagnoses (**Table 3**). The three MCAS-associated MC symptoms with the highest percentage in hEDS/HSD patients with Hi MC scores included headache (hEDS OR 5.74, CI 3.74-8.72, *p* = 0.0001; HSD OR 4.63, CI 3.09-6.99, *p* < 0.0001), nasal congestion (hEDS OR 42.20, CI 18.31-90.32, *p* < 0.0001; HSD OR 12.41, CI 8.33-18.14, *p* < 0.0001), and migraine (hEDS OR 3.72, CI 2.14-6.44, *p* < 0.0001; HSD OR 4.14, CI 2.93-5.81, *p* < 0.0001) (**Table 3**, **Supplementary Table 3**, **Fig1d,e**). The third most reported symptom was dizziness, but that was reported more often only in patients with HSD with a Hi MC score (HSD OR 2.74, CI 1.92-3.92, *p* < 0.0001) (**Table 3**). Thus, hEDS/HSD patients with high MC scores reported more MCAS-related symptoms/comorbidities than patients with low MC scores. Furthermore, these findings indicate that overlap exists between the MCAS symptoms/comorbidities reported by Akin and Valent studies and a high MC score.

**Table 3.** MCAS-related symptoms/comorbidities from Akin et al.^13^ and/or Valent et al.^14^ in patients diagnosed with hEDS or HSD with low (Lo) vs. high (Hi) MC scores (*n* = 1,050)

| Condition | hEDS<sup>a</sup><br>Lo MC<br>($n = 98$) | hEDS<br>Hi MC<br>($n = 133$) | *P* value<sup>b</sup> | HSD<br>Lo MC<br>($n = 294$) | HSD<br>Hi MC<sup>c</sup><br>($n = 328$) | *P* value<sup>b</sup> |
| --- | --- | --- | --- | --- | --- | --- |
| Headache | 66 (67.3) | 118 (88.7) | **0.0001<sup>d</sup>** | 179 (60.9) | 288 (87.8) | **<0.0001** |
| Nasal congestion | 34 (34.7) | 120 (90.2) | **<0.0001** | 94 (32.0) | 280 (85.4) | **<0.0001** |
| Dizzy | 72 (73.5) | 110 (82.7) | 0.10 | 174 (59.2) | 262 (79.9) | **<0.0001** |
| Migraine | 44 (44.9) | 100 (75.2) | **<0.0001** | 132 (44.9) | 253 (77.1) | **<0.0001** |
| Nausea | $n = 115$ | $n = 144$ | **0.0005** | $n = 400$ | $n = 391$ | **<0.0001** |
|  | 63 (54.8) | 109 (75.7) |  | 220 (55.0) | 295 (75.5) |  |
| Pruritus (itching) | 30 (30.6) | 102 (76.7) | **<0.0001** | 77 (26.2) | 240 (73.2) | **<0.0001** |
| Abdominal cramping | 63 (54.8) | 102 (70.8) | **0.009** | 204 (51.0) | 274 (70.3) | **<0.0001** |
| Flushing | 28 (28.6) | 104 (78.2) | **<0.0001** | 91 (31.0) | 219 (66.8) | **<0.0001** |
| Tachycardia | 45 (45.9) | 92 (69.2) | **0.0004** | 113 (38.4) | 210 (64.0) | **<0.0001** |
| Urticaria (hives) | 17 (17.3) | 92 (69.2) | **<0.0001** | 39 (13.3) | 198 (60.4) | **<0.0001** |
| Diarrhea | 35 (35.7) | 85 (63.9) | **<0.0001** | 81 (27.6) | 183 (55.8) | **<0.0001** |
| Difficulty breathing | 24 (24.5) | 73 (54.9) | **<0.0001** | 45 (15.3) | 150 (45.7) | **<0.0001** |
| Throat swelling | 10 (10.2) | 80 (60.2) | **<0.0001** | 33 (11.2) | 138 (42.1) | **<0.0001** |
| Hypotension | 23 (23.5) | 64 (48.1) | **0.0002** | 48 (16.3) | 123 (37.5) | **<0.0001** |
| Vomiting | 21 (21.4) | 46 (34.6) | **0.040** | 39 (13.3) | 110 (33.5) | **<0.0001** |
| Wheezing | $n = 115$ | $n = 144$ | **<0.0001** | $n = 400$ | $n = 390$ | **<0.0001** |
|  | 3 (2.6) | 36 (25.0) |  | 20 (5.0) | 98 (25.1) |  |
| Weal/blisters | 4 (4.1) | 34 (25.6) | **<0.0001** | 15 (5.1%) | 79 (24.1) | **<0.0001** |
| Collapse | 19 (19.4) | 42 (31.6) | **0.049** | 44 (15.0) | 74 (22.6) | **0.018** |
| None of the above | 17 (17.3) | 3 (2.3) | **<0.0001** | 61 (20.7) | 4 (1.2) | **<0.0001** |

<sup>a</sup> All data shown as $n$ (%). <sup>b</sup> *P* values obtained from Fisher's exact test for categorical data. <sup>c</sup> Order of conditions based on highest to lowest percentage in HSD Hi MC group. <sup>d</sup> Bold indicates significant value $p < 0.05$ ; values higher than $p < 0.0004$ were not significant after Bonferroni correction.

### 3.4 Blood and urinary MC markers

To determine whether a high MC score related to greater blood and urinary markers of MC activation, we examined randomly collected blood and urine samples from patients seen at the EDS Clinic that were diagnosed with hEDS or HSD. We found that patients with hEDS (*p* = 0.93) or HSD (*p* = 0.14) with a Hi MC score did not have significantly higher blood tryptase levels than patients with a Lo MC score (**Fig2a**). Tryptase is a product of MC degranulation and used as part of the diagnostic criteria for MCAS.^14^ However, we did not determine the ‘baseline’ level of tryptase for individual patients in order to determine whether their tryptase level was above this level as required for a MCAS diagnosis.

**Figure 2.**
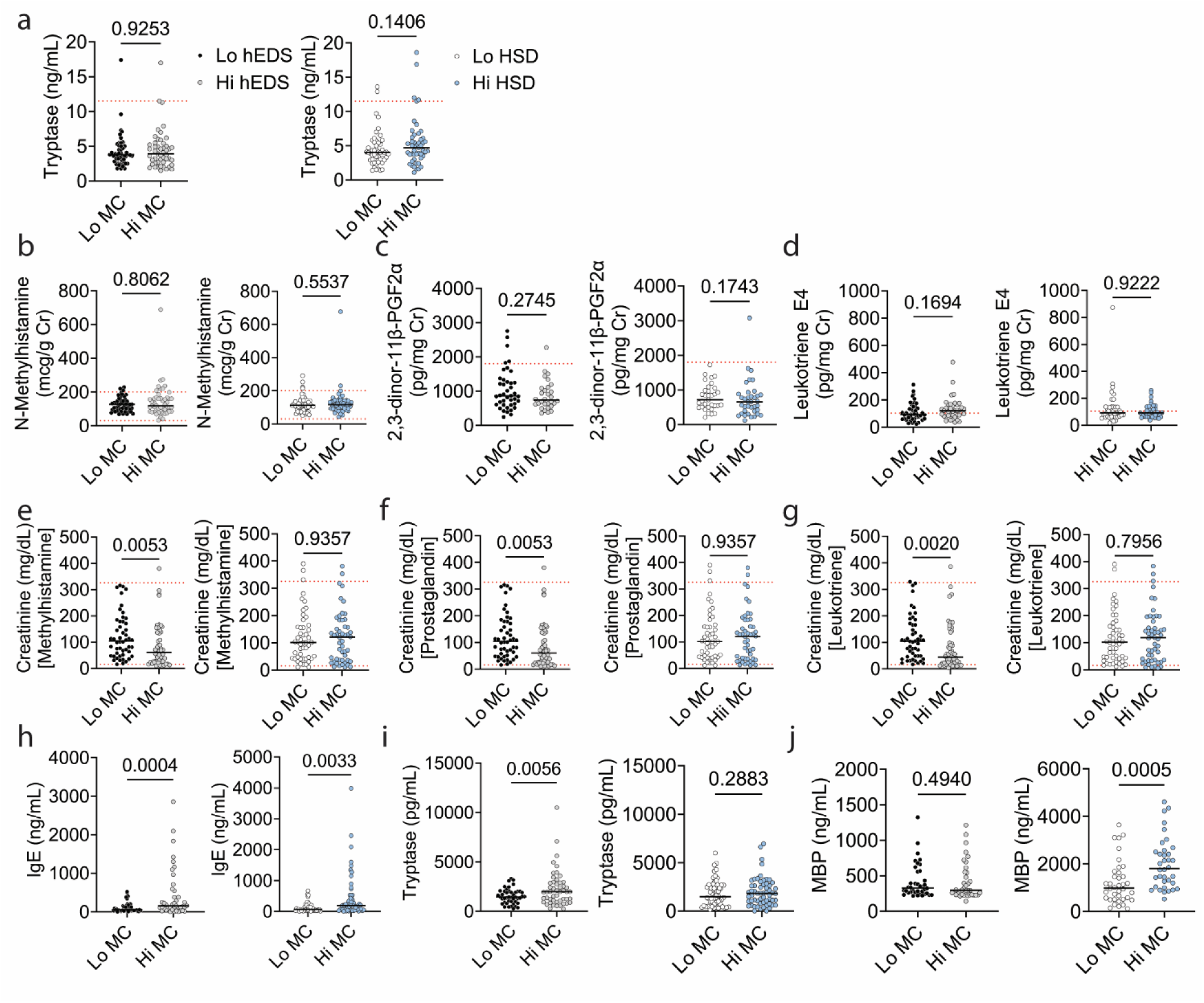
Blood and urine MC markers. Random clinically tested samples of MC-related metabolites of hEDS (black/grey) and HSD (white/blue) patients comparing low (Lo) vs. high (Hi) MC scores for a) blood tryptase (ng/mL) b) N-methylhistamine (μg/g creatinine/Cr), c) 2,3-donor-11β-prostaglandin/PG F2α (pg/μg Cr), and d) leukotriene E4 (pg/mg Cr). e-g) Urine creatinine (Cr) levels for each test. Red lines indicate upper and lower limits for normal values. ELISA of random banked blood samples for h) total IgE (ng/mL), i) tryptase (pg/mL) and j) major basic protein (MBP) (ng/mL). a-g) *n* = 50/group, h) *n* = 42-58/group, i) *n* = 50/group, j) *n* = 38/group. P values derived from Student’s *t* test for parametric data and Mann-Whitney rank test for nonparametric data.

Similarly, we did not find an increase based on MC score for hEDS/HSD in urinary levels of the MC metabolites N-methylhistamine (hEDS *p* = 0.81, HSD *p* = 0.55) (**Fig2b**), 2,3-dinor-11β-prostaglandin F2α (PGF) (hEDS *p* = 0.28, HSD *p* = 0.17) (**Fig2c**), or leukotriene E4 (hEDS *p* = 0.17, HSD *p* = 0.92) (**Fig2d**) processed at the Mayo Clinic Clinical Laboratory. Because of differences in hydration between patients, urine protein levels are normalized to urinary creatinine (Cr) levels. Interestingly, we found that patients with hEDS and a high MC score had significantly less urinary creatinine than hEDS patients with Lo MC scores for each of the urinary MC metabolites that we examined (creatinine used for the N-methylhistamine test *p* = 0.005, creatinine used for the PGF test *p* = 0.005, and creatinine used for the leukotriene test *p* = 0.002) (**Fig2e-g**). These findings indicate that the random urinary MC metabolite tests for hEDS patients were not accurate because the protein used to normalize the values was altered by the analysis parameters.

In a separate set of patients, we examined several MC markers in the blood of banked samples that were collected on the same day as patients were seen at the EDS Clinic (i.e., random samples). We found that patients with hEDS (*p* = 0.0004) or HSD (*p* = 0.003) and Hi MC scores had significantly increased total IgE levels in the blood by ELISA compared to patients with Lo MC scores (**Fig2h**). IgE is a MC-specific marker. Similarly, we found that hEDS patients with a high MC score had increased tryptase levels compared to patients with Lo MC scores (*p* = 0.006) (**Fig2i**). But this was not found for HSD patients (*p* = 0.29) (**Fig2i**). This tryptase ELISA was processed in the Fairweather lab. In contrast, when we examined major basic protein (MBP) levels in the blood, we found that patients with HSD and a high MC score had elevated levels of MBP compared to patients with Lo MC scores (*p* = 0.0005) (**Fig2j**). MBP is a mediator released from eosinophils, which are activated by MCs. Thus, randomly collected clinical blood and urinary measurements of MC activation were not related to a high MC score but other blood MC markers were found to associate with high MC scores including IgE, tryptase (Fairweather lab ELISA) and MBP.

### 3.5 Skin conditions

Next, we examined whether a high MC score was associated with more self-reported symptoms and comorbidities from the Intake Questionnaire. Note that differences in percentages between groups are shown in the Tables while ORs and CIs are shown in Supplementary Tables for all symptoms/comorbidities. We found that hEDS/HSD patients with a Hi MC score reported more skin conditions than those with a Lo MC score including issues that are part of the diagnostic criteria for hEDS^4^ such as easy bruising (hEDS OR 3.16, CI 1.39-7.51, *p* < 0.006; HSD OR 2.01, CI 1.36-2.99, *p* < 0.0005), easy scarring (hEDS OR 2.05, CI 1.05-3.97, *p* < 0.009; HSD OR 2.05, CI 1.49-2.81, *p* < 0.0001), stretch marks (hEDS OR 1.94, CI 1.12-3.30, *p* < 0.025; HSD OR 1.96, CI 1.46-2.62, *p* < 0.0001), poor wound healing (hEDS OR 3.01, CI 1.73-5.25, *p* < 0.0001; HSD OR 2.26, CI 1.69-3.01, *p* < 0.0001), and abnormal scarring (hEDS OR 2.50, CI 1.49-4.15, *p* < 0.0001; HSD OR 3.03, CI 2.26-4.05, *p* < 0.0001) (**Table 4**, **Supplementary Table 4**). Other skin issues such as eczema were also reported more often in hEDS/HSD patients with Hi MC activity: (hEDS OR 20.30, CI 8.63-45.61, *p* < 0.0001; HSD OR 10.95, CI 7.25-16.48, *p* < 0.0001) (**Table 4**, **Supplementary Table 4**). Thus, many of the skin conditions that we examined were reported more frequently in hEDS/HSD patients with a high MC score.

**Table 4.** Skin conditions in patients diagnosed with hEDS or HSD with low (Lo) vs. high (Hi) MC scores (*n* = 1,050)

| Condition | hEDS<sup>a</sup><br>Lo MC<br>($n = 115$) | hEDS<br>Hi MC<br>($n = 144$) | *P* value<sup>b</sup> | HSD<br>Lo MC<br>($n = 400$) | HSD<sup>c</sup><br>Hi MC<br>($n = 391$) | *P* value<sup>b</sup> |
| --- | --- | --- | --- | --- | --- | --- |
| Easy bruising | 95 (82.6) | 135 (93.8) | **0.006<sup>d</sup>** | 317 (79.3) | 346 (88.5) | **0.0005** |
| Easy scarring | 89 (77.4) | 126 (87.5) | **0.045** | 255 (63.8) | 306 (78.3) | **<0.0001** |
| Stretch marks | 75 (65.2) | 113 (78.5) | **0.025** | 229 (57.3) | 283 (72.4) | **<0.0001** |
| Poor wound healing | 85 (57.4) | 173 (80.5) | **<0.0001** | 224 (49.3) | 320 (67.4) | **<0.0001** |
| Abnormal scarring | 46 (40.0) | 90 (62.5) | **0.0004** | 110 (27.5) | 209 (53.5) | **<0.0001** |
| Eczema | 6 (5.2) | 76 (52.8) | **<0.0001** | 33 (8.3) | 194 (49.6) | **<0.0001** |
| Rosacea | 15 (13.0) | 14 (9.7) | 0.43 | 39 (9.8) | 86 (21.9) | **<0.0001** |
| Psoriasis | 9 (7.8) | 21 (14.6) | 0.12 | 14 (3.5) | 54 (13.8) | **<0.0001** |
| Other | 13 (11.3) | 35 (24.3) | **0.010** | 40 (10.0) | 97 (24.8) | **<0.0001** |
| No issues | 52 (45.2) | 14 (9.7) | **<0.0001** | 194 (48.5) | 50 (12.8) | **<0.0001** |
| Unknown | 25 (21.7) | 24 (16.7) | 0.34 | 91 (22.8) | 58 (14.8) | **0.005** |

<sup>a</sup> All data shown as $n$ (%). <sup>b</sup> *P* values obtained from Fisher's exact test for categorical data. <sup>c</sup> Order of conditions based on highest to lowest percentage in HSD Hi MC group. <sup>d</sup> Bold indicates significant value $p < 0.05$ ; values higher than $p < 0.0004$ were not significant after Bonferroni correction.

### 3.6 Joint issues

We found that hEDS/HSD patients with Hi MC scores self-reported more joint issues than patients with Lo MC scores including joint pain (hEDS OR 4.27, CI 1.69-10.72, *p* = 0.002, HSD OR 4.02, CI 1.91-8.53, *p* = 0.0002), joint subluxations (hEDS OR 4.43, CI 2.15-8.82, *p* < 0.0001, HSD OR 2.33, CI 1.63-3.30, *p* < 0.0001), joint sprains (hEDS OR 4.38, CI 2.36-7.93, *p* < 0.0001, HSD OR 3.77, C 2.72-5.22, *p* < 0.0001), and joint dislocations (hEDS OR 2.38, CI 1.44-3.95, *p* = 0.001, HSD OR 1.87, CI 1.37-2.54, *p* < 0.0001) (**Table 5**, **Supplementary Table 5**). Thus, patients with high MC scores reported worse joint symptoms.

**Table 5.** Joint issues in patients diagnosed with hEDS or HSD with low (Lo) vs. high (Hi) MC scores (*n* = 1,050)

| Condition | hEDS<sup>a</sup><br>Lo MC<br>($n = 115$) | hEDS<br>Hi MC<br>($n = 144$) | *P* value<sup>b</sup> | HSD<br>Lo MC<br>($n = 400$) | HSD<sup>c</sup><br>Hi MC<br>($n = 391$) | *P* value<sup>b</sup> |
| --- | --- | --- | --- | --- | --- | --- |
| Joint pain | 97 (84.4) | 138 (95.8) | **0.002<sup>c</sup>** | 369 (92.3) | 383 (97.9) | **0.0002** |
| Joint subluxations | 82 (71.3) | 132 (91.7) | **<0.0001** | 283 (70.8) | 332 (84.9) | **<0.0001** |
| Joint sprains | 71 (61.7) | 126 (87.5) | **<0.0001** | 216 (54.0) | 319 (81.6) | **<0.0001** |
| Joint dislocations | 34 (29.6) | 72 (50.0) | **0.001** | 100 (25.0) | 150 (38.4) | **<0.0001** |
| None | 6 (5.2) | 0 (0.0) | **0.007** | 11 (2.8) | 1 (0.3) | **0.006** |

<sup>a</sup> All data shown as $n$ (%). <sup>b</sup> *P* values obtained from Fisher's exact test for categorical data. <sup>c</sup> Order of conditions based on highest to lowest percentage in HSD Hi MC group. <sup>d</sup> Bold indicates significant value $p < 0.05$ ; values higher than $p < 0.0004$ were not significant after Bonferroni correction.

### 3.7 Gastrointestinal symptoms and comorbidities

When we examined gastrointestinal (GI) symptoms/comorbidities, we found that hEDS and HSD patients with a Hi MC score reported more GI symptoms than those with Lo MC scores for most of the conditions that we examined (**Table 6**). The top 5 symptoms/comorbidities for hEDS/HSD included nausea (hEDS OR 2.57, CI 1.51-4.35, *p* = 0.0005, HSD OR 2.51, CI 1.85-3.41, *p* < 0.0001), constipation (hEDS OR 2.86, CI 1.67-4.79, *p* < 0.0001, HSD OR 2.44, CI 1.83-3.27, *p* < 0.0001), diarrhea (hEDS OR 2.40, CI 1.46-4.02, *p* = 0.0009, HSD OR 2.78, CI 2.08-3.70, *p* < 0.0001), gastroesophageal reflux disease (GERD) (hEDS OR 3.75, CI 2.21-6.45, *p* < 0.0001, HSD OR 2.95, CI 2.20-3.96, *p* < 0.0001), and irritable bowel syndrome (IBS) (hEDS OR 2.86, CI 1.67-4.79, *p* < 0.0001, HSD OR 2.83, CI 2.11-3.81, *p* < 0.0001) (**Table 6**, **Supplemental Table 6**). Thus, more hEDS/HSD patients with a high MC score reported GI symptoms and comorbidities.

**Table 6.** Gastrointestinal symptoms in patients diagnosed with hEDS or HSD with low (Lo) vs. high (Hi) MC scores (*n* = 1,050)

| Condition | hEDS<sup>a</sup><br>Lo MC<br>($n = 115$) | hEDS<br>Hi MC<br>($n = 144$) | *P* value<sup>b</sup> | HSD<br>Lo MC<br>($n = 400$) | HSD<sup>c</sup><br>Hi MC<br>($n = 391$) | *P* value<sup>b</sup> |
| --- | --- | --- | --- | --- | --- | --- |
| Nausea | 63 (54.8) | 109 (75.7) | **0.0005<sup>d</sup>** | 220 (55.0) | 295 (75.5) | **<0.0001** |
| Constipation | 58 (50.4) | 108 (75.0) | **<0.0001** | 207 (51.8) | 283 (72.4) | **<0.0001** |
| Diarrhea | 55 (47.8) | 99 (68.8) | **0.0009** | 169 (42.3) | 262 (67.0) | **<0.0001** |
| GERD<sup>e</sup> | 30 (26.1) | 82 (56.9) | **<0.0001** | 124 (31.0) | 223 (57.0) | **<0.0001** |
| IBS | 28 (24.4) | 69 (47.9) | **0.0001** | 104 (26.0) | 195 (49.9) | **0.0001** |
| Dyspepsia | 20 (17.4) | 60 (41.7) | **<0.0001** | 98 (24.5) | 183 (46.8) | **<0.0001** |
| Vomiting | 32 (27.8) | 67 (46.5) | **0.003** | 99 (24.8) | 173 (44.3) | **<0.0001** |
| Hemorrhoids | 28 (24.4) | 70 (48.6) | **<0.0001** | 85 (21.3) | 150 (38.4) | **<0.0001** |
| Gastroparesis | 13/ (11.3) | 44 (30.6) | **0.0002** | 65 (16.3) | 105 (26.9) | **0.0004** |
| Hernias | 19 (16.5) | 44 (30.6) | **0.0091** | 48 (12.0) | 103 (26.3) | **<0.0001** |
| Anal fissure | 14 (12.2) | 36 (25.0) | **0.011** | 27 (6.8) | 73 (18.7) | **<0.0001** |
| Fecal incontinence | 3 (2.6) | 15 (10.4) | **0.014** | 16 (4.0) | 28 (7.2) | 0.06 |
| Rectal prolapse | 7 (6.1) | 16 (11.1) | 0.19 | 11 (2.8) | 24 (6.1) | **0.024** |
| Barrett's esophagus | 1 (0.9) | 5 (3.5) | 0.23 | 2 (0.5) | 10 (2.6) | **0.020** |
| Ulcerative colitis | 4 (3.5) | 7 (4.9) | 0.76 | 3 (0.8) | 8 (2.1) | 0.14 |
| Crohn's disease | 1 (0.9) | 2 (1.4) | 0.99 | 2 (0.5) | 6 (1.5) | 0.17 |
| Other | 12 (10.4) | 39 (27.1) | **0.0009** | 42 (10.5) | 74 (18.9) | **0.0009** |
| No issues | 12 (10.4) | 0 (0.0) | **<0.0001** | 44 (11.0) | 4 (1.0) | **<0.0001** |
| Unknown | 8 (6.9) | 1 (0.7) | **0.012** | 12 (3.0) | 4 (1.0) | 0.07 |

<sup>a</sup> All data shown as $n$ (%). <sup>b</sup> *P* values obtained from Fisher's exact test for categorical data. <sup>c</sup> Order of conditions based on highest to lowest percentage in HSD Hi MC group. <sup>d</sup> Bold indicates significant value $p < 0.05$ ; values higher than $p < 0.0004$ were not significant after Bonferroni correction. <sup>e</sup> Abbreviations: GERD, gastroesophageal reflux disease; IBS, irritable bowel syndrome.

### 3.8 Genitourinary symptoms and comorbidities

When we examined genitourinary issues, we found that hEDS and HSD patients with a Hi MC score reported more genitourinary symptoms than those with Lo MC scores for many of the conditions that we examined (**Table 7**). The top 5 symptoms/comorbidities for hEDS/HSD included dyspareunia (persistent or recurrent pain during sexual intercourse) (hEDS OR 3.39, CI 1.92-5.96, *p* < 0.0001; HSD OR 2.63, CI 1.90-3.64, *p* < 0.0001), recurrent urinary tract infection (hEDS OR 2.56, CI 1.49-4.52, *p* = 0.001; HSD OR 2.06, CI 1.50-2.84, *p* < 0.0001), incontinence (hEDS OR 2.45, CI 1.32-4.58, *p* = 0.005; HSD OR 2.72, CI 1.93-3.79, *p* < 0.0001), pelvic floor dysfunction (hEDS OR 3.84, CI 1.99-7.29, *p* < 0.0001; HSD OR 3.45, CI 2.32-5.21, *p* < 0.0001), and polycystic ovary syndrome (PCOS) (hEDS OR 3.50, CI 1.64-7.77, *p* = 0.001; HSD OR 2.23, CI 1.54-3.26, *p* < 0.0001) (**Table 7**, **Supplementary Table 7**). Thus, more patients with high MC scores reported genitourinary issues.

**Table 7.** Genitourinary symptoms in patients diagnosed with hEDS or HSD with low (Lo) vs. high (Hi) MC scores (*n* = 1,050)

| Condition | hEDS<sup>a</sup><br>Lo MC<br>($n = 115$) | hEDS<br>Hi MC<br>($n = 144$) | *P* value<sup>b</sup> | HSD<br>Lo MC<br>($n = 400$) | HSD<sup>c</sup><br>Hi MC<br>($n = 391$) | *P* value<sup>b</sup> |
| --- | --- | --- | --- | --- | --- | --- |
| Dyspareunia | 24 (20.9) | 68 (47.2) | **<0.0001<sup>d</sup>** | 74 (18.5) | 146 (37.3) | **<0.0001** |
| Recurrent urinary tract infection | 24 (20.9) | 58 (40.3) | **0.001** | 80 (20.0) | 133 (34.0) | **<0.0001** |
| Incontinence | 17 (14.8) | 43 (29.9) | **0.005** | 62 (15.5) | 130 (33.2) | **<0.0001** |
| Pelvic floor dysfunction | 14 (12.2) | 50 (34.7) | **<0.0001** | 38 (9.5) | 104 (26.6) | **<0.0001** |
| PCOS<sup>e</sup> | 9 (7.8) | 33 (22.9) | **0.001** | 51 (12.8) | 96 (24.6) | **<0.0001** |
| Recurrent yeast infection | 13 (11.3) | 31 (21.5) | **0.031** | 45 (11.3) | 95 (24.3) | **<0.0001** |
| Endometriosis | 15 (13.0) | 45 (31.3) | **0.0006** | 42 (10.5) | 95 (24.3) | **<0.0001** |
| Pelvic floor spasm | 11 (9.6) | 27 (18.8) | 0.05 | 19 (4.8) | 70 (17.9) | **<0.0001** |
| Interstitial cystitis | 7 (6.1) | 20 (13.9) | **0.043** | 12 (3.0) | 56 (14.3) | **<0.0001** |
| Recurrent vaginal bacterial infection | 10 (8.7) | 15 (10.4) | 0.68 | 25 (6.3) | 45 (11.5) | **0.012** |
| Uterine prolapse | 5 (4.3) | 13 (9.0) | 0.22 | 10 (2.5) | 22 (5.6) | **0.030** |
| Bladder prolapse | 5 (4.3) | 14 (9.7) | 0.15 | 8 (2.0) | 22 (5.6) | **0.009** |
| Other | 10 (8.7) | 25 (17.4) | **0.046** | 44 (11.0) | 66 (16.9) | **0.018** |
| No issues | 42 (36.5) | 18 (12.5) | **<0.0001** | 127 (31.8) | 41 (10.5) | **<0.0001** |
| Unknown | 14 (12.2) | 9 (6.3) | 0.12 | 54 (13.5) | 31 (7.9) | **0.012** |

<sup>a</sup> All data shown as $n$ (%). <sup>b</sup> *P* values obtained from Fisher's exact test for categorical data. <sup>c</sup> Order of conditions based on highest to lowest percentage in HSD Hi MC group. <sup>d</sup> Bold indicates significant value $p < 0.05$ ; values higher than $p < 0.0004$ were not significant after Bonferroni correction. <sup>e</sup> Abbreviations: PCOS, polycystic ovary syndrome.

### 3.8 Neurological symptoms and comorbidities

When we examined neurological symptoms/comorbidities, we found that hEDS/HSD patients with a Hi MC score reported more neurological symptoms than those with Lo MC score for most of the conditions that we examined (**Table 8**). The top 5 neurological symptoms were brain fog (hEDS OR 3.56, CI 2.25-5.53, *p* = 0.00013; HSD OR 2.15 CI 1.51-3.08, *p* < 0.0001), headache (hEDS OR 1.39, CI 0.73-2.64; *p* = 0.33, EDS OR 2.05, CI 1.46-2.84, *p* < 0.0001), migraine (hEDS OR 2.94, CI 1.78-4.82, *p* < 0.0001; HSD OR 3.57, CI 2.65-4.82, *p* < 0.0001), tinnitus (hEDS OR 3.67, CI 2.21-6.06, *p* < 0.0001; HSD OR 1.50, CI 1.13-1.98, *p* < 0.0001), and vertigo (hEDS OR 2.32, CI 1.41-3.80, *p* = 0.002, HSD OR 2.35, CI 1.74-3.16, *p* < 0.0001) (**Table 8**, **Supplementary Table 8**). Thus, more hEDS/HSD patients with a high MC score report neurological symptoms and comorbidities.

**Table 8.**
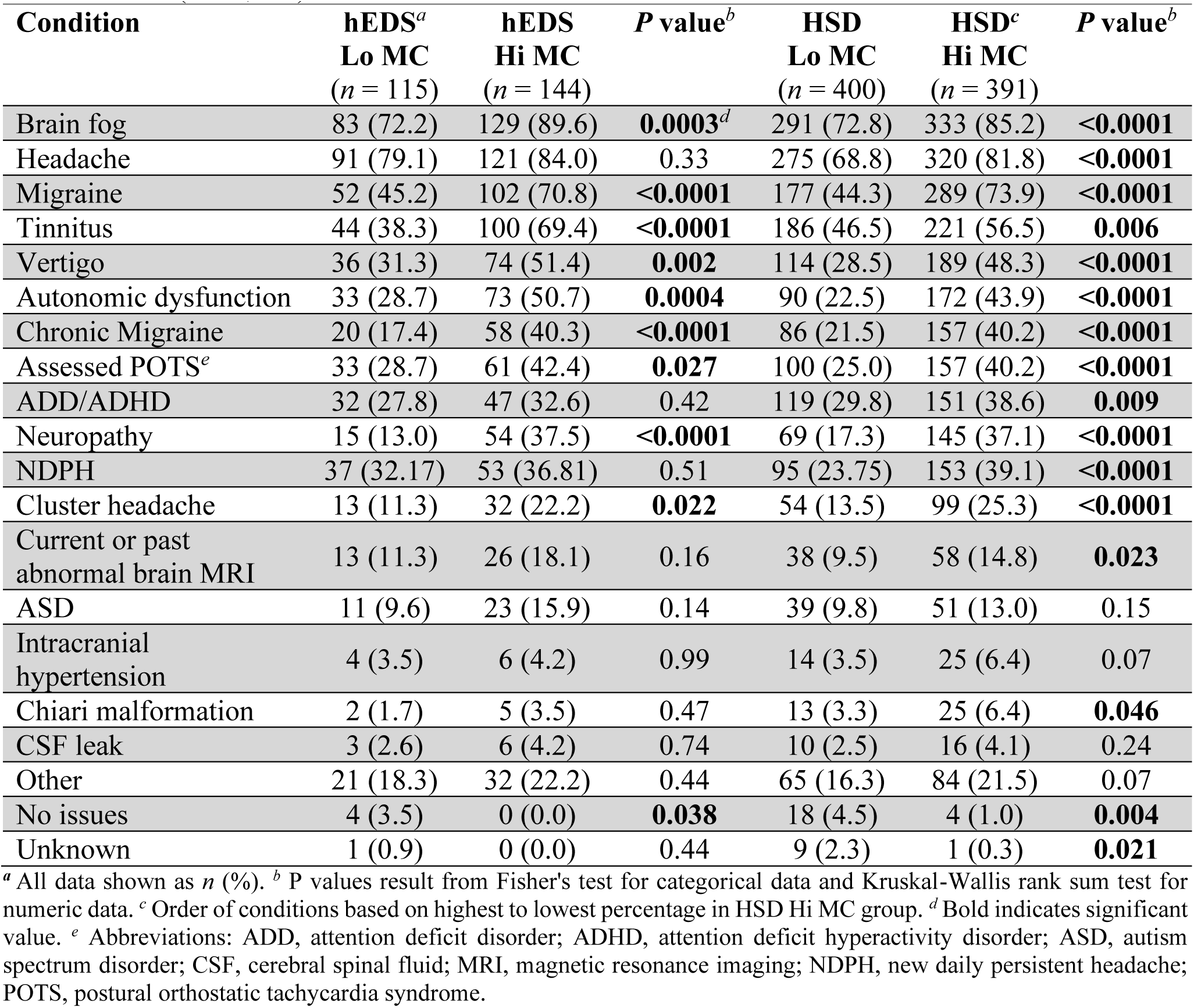
Neurological conditions in patients with low (Lo) vs. high (Hi) MC scores diagnosed with hEDS or HSD (*n* = 1,050)

| Condition | hEDS<sup>a</sup><br>Lo MC<br>($n = 115$) | hEDS<br>Hi MC<br>($n = 144$) | *P* value<sup>b</sup> | HSD<br>Lo MC<br>($n = 400$) | HSD<sup>c</sup><br>Hi MC<br>($n = 391$) | *P* value<sup>b</sup> |
| --- | --- | --- | --- | --- | --- | --- |
| Brain fog | 83 (72.2) | 129 (89.6) | **0.0003<sup>d</sup>** | 291 (72.8) | 333 (85.2) | **<0.0001** |
| Headache | 91 (79.1) | 121 (84.0) | 0.33 | 275 (68.8) | 320 (81.8) | **<0.0001** |
| Migraine | 52 (45.2) | 102 (70.8) | **<0.0001** | 177 (44.3) | 289 (73.9) | **<0.0001** |
| Tinnitus | 44 (38.3) | 100 (69.4) | **<0.0001** | 186 (46.5) | 221 (56.5) | **0.006** |
| Vertigo | 36 (31.3) | 74 (51.4) | **0.002** | 114 (28.5) | 189 (48.3) | **<0.0001** |
| Autonomic dysfunction | 33 (28.7) | 73 (50.7) | **0.0004** | 90 (22.5) | 172 (43.9) | **<0.0001** |
| Chronic Migraine | 20 (17.4) | 58 (40.3) | **<0.0001** | 86 (21.5) | 157 (40.2) | **<0.0001** |
| Assessed POTS<sup>e</sup> | 33 (28.7) | 61 (42.4) | **0.027** | 100 (25.0) | 157 (40.2) | **<0.0001** |
| ADD/ADHD | 32 (27.8) | 47 (32.6) | 0.42 | 119 (29.8) | 151 (38.6) | **0.009** |
| Neuropathy | 15 (13.0) | 54 (37.5) | **<0.0001** | 69 (17.3) | 145 (37.1) | **<0.0001** |
| NDPH | 37 (32.17) | 53 (36.81) | 0.51 | 95 (23.75) | 153 (39.1) | **<0.0001** |
| Cluster headache | 13 (11.3) | 32 (22.2) | **0.022** | 54 (13.5) | 99 (25.3) | **<0.0001** |
| Current or past<br>abnormal brain MRI | 13 (11.3) | 26 (18.1) | 0.16 | 38 (9.5) | 58 (14.8) | **0.023** |
| ASD | 11 (9.6) | 23 (15.9) | 0.14 | 39 (9.8) | 51 (13.0) | 0.15 |
| Intracranial<br>hypertension | 4 (3.5) | 6 (4.2) | 0.99 | 14 (3.5) | 25 (6.4) | 0.07 |
| Chiari malformation | 2 (1.7) | 5 (3.5) | 0.47 | 13 (3.3) | 25 (6.4) | **0.046** |
| CSF leak | 3 (2.6) | 6 (4.2) | 0.74 | 10 (2.5) | 16 (4.1) | 0.24 |
| Other | 21 (18.3) | 32 (22.2) | 0.44 | 65 (16.3) | 84 (21.5) | 0.07 |
| No issues | 4 (3.5) | 0 (0.0) | **0.038** | 18 (4.5) | 4 (1.0) | **0.004** |
| Unknown | 1 (0.9) | 0 (0.0) | 0.44 | 9 (2.3) | 1 (0.3) | **0.021** |

<sup>a</sup> All data shown as $n$ (%). <sup>b</sup> *P* values result from Fisher's test for categorical data and Kruskal-Wallis rank sum test for numeric data. <sup>c</sup> Order of conditions based on highest to lowest percentage in HSD Hi MC group. <sup>d</sup> Bold indicates significant value. <sup>e</sup> Abbreviations: ADD, attention deficit disorder; ADHD, attention deficit hyperactivity disorder; ASD, autism spectrum disorder; CSF, cerebral spinal fluid; MRI, magnetic resonance imaging; NDPH, new daily persistent headache; POTS, postural orthostatic tachycardia syndrome.

### 3.9 Fibromyalgia symptoms

The Intake Questionnaire contains the questions used to diagnose fibromyalgia. For a formal diagnosis patients are additionally assessed during their in-person appointment, according to the diagnostic criteria for fibromyalgia.^34^ In Table 9 we list the fibromyalgia diagnosis based only on the questionnaire. Table 9 also contains the percentage of patients that answered each of the questions for symptoms that comprise the fibromyalgia assessment.

**Table 9.**
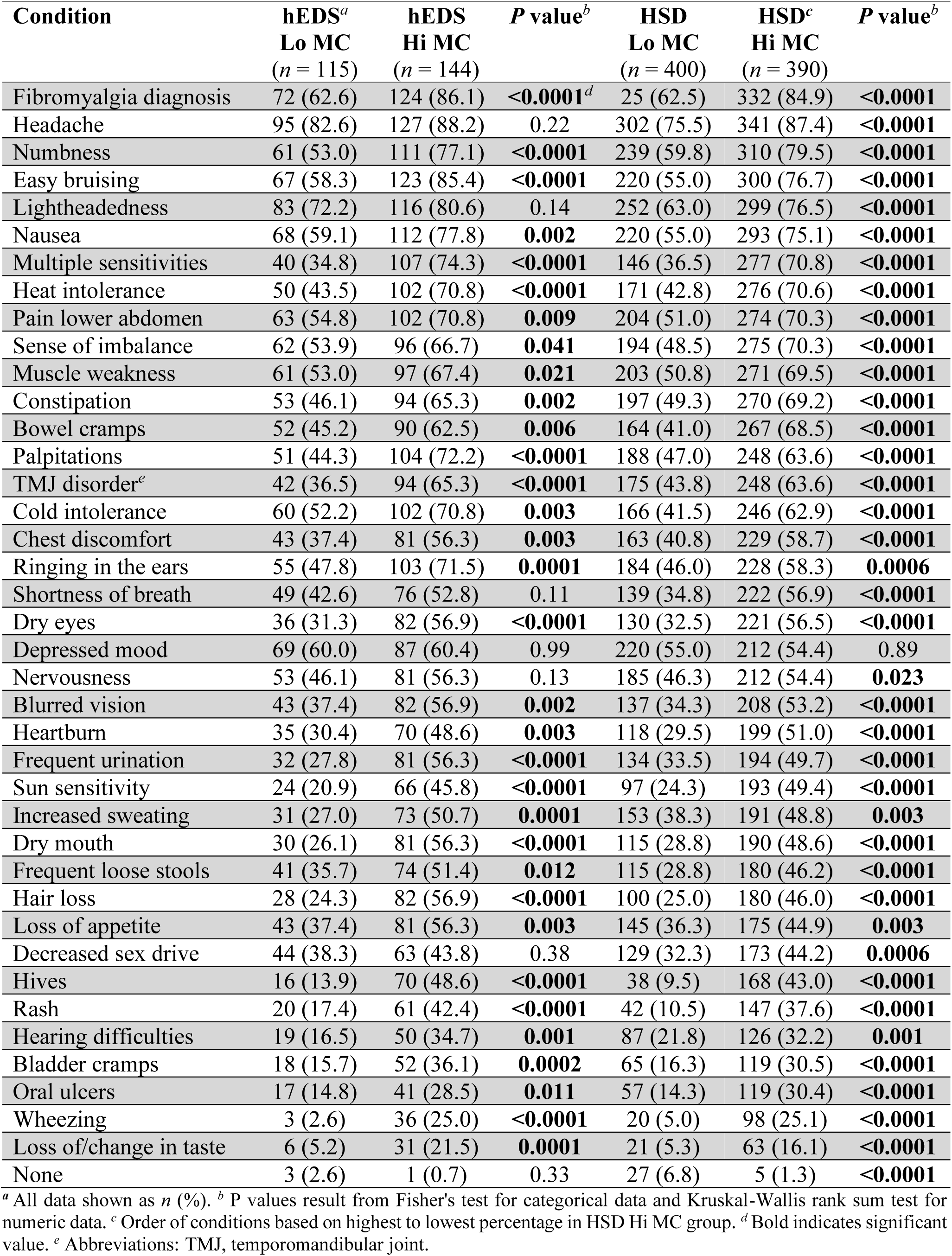
Fibromyalgia diagnosis and fibromyalgia-associated symptoms in patients diagnosed with hEDS or HSD with low (Lo) vs. high (Hi) MC scores (*n* = 1,050)

| Condition | hEDS<sup>a</sup><br>Lo MC<br>($n = 115$) | hEDS<br>Hi MC<br>($n = 144$) | $P$ value<sup>b</sup> | HSD<br>Lo MC<br>($n = 400$) | HSD<sup>c</sup><br>Hi MC<br>($n = 390$) | $P$ value<sup>b</sup> |
| --- | --- | --- | --- | --- | --- | --- |
| Fibromyalgia diagnosis | 72 (62.6) | 124 (86.1) | **<0.0001<sup>d</sup>** | 25 (62.5) | 332 (84.9) | **<0.0001** |
| Headache | 95 (82.6) | 127 (88.2) | 0.22 | 302 (75.5) | 341 (87.4) | **<0.0001** |
| Numbness | 61 (53.0) | 111 (77.1) | **<0.0001** | 239 (59.8) | 310 (79.5) | **<0.0001** |
| Easy bruising | 67 (58.3) | 123 (85.4) | **<0.0001** | 220 (55.0) | 300 (76.7) | **<0.0001** |
| Lightheadedness | 83 (72.2) | 116 (80.6) | 0.14 | 252 (63.0) | 299 (76.5) | **<0.0001** |
| Nausea | 68 (59.1) | 112 (77.8) | **0.002** | 220 (55.0) | 293 (75.1) | **<0.0001** |
| Multiple sensitivities | 40 (34.8) | 107 (74.3) | **<0.0001** | 146 (36.5) | 277 (70.8) | **<0.0001** |
| Heat intolerance | 50 (43.5) | 102 (70.8) | **<0.0001** | 171 (42.8) | 276 (70.6) | **<0.0001** |
| Pain lower abdomen | 63 (54.8) | 102 (70.8) | **0.009** | 204 (51.0) | 274 (70.3) | **<0.0001** |
| Sense of imbalance | 62 (53.9) | 96 (66.7) | **0.041** | 194 (48.5) | 275 (70.3) | **<0.0001** |
| Muscle weakness | 61 (53.0) | 97 (67.4) | **0.021** | 203 (50.8) | 271 (69.5) | **<0.0001** |
| Constipation | 53 (46.1) | 94 (65.3) | **0.002** | 197 (49.3) | 270 (69.2) | **<0.0001** |
| Bowel cramps | 52 (45.2) | 90 (62.5) | **0.006** | 164 (41.0) | 267 (68.5) | **<0.0001** |
| Palpitations | 51 (44.3) | 104 (72.2) | **<0.0001** | 188 (47.0) | 248 (63.6) | **<0.0001** |
| TMJ disorder<sup>e</sup> | 42 (36.5) | 94 (65.3) | **<0.0001** | 175 (43.8) | 248 (63.6) | **<0.0001** |
| Cold intolerance | 60 (52.2) | 102 (70.8) | **0.003** | 166 (41.5) | 246 (62.9) | **<0.0001** |
| Chest discomfort | 43 (37.4) | 81 (56.3) | **0.003** | 163 (40.8) | 229 (58.7) | **<0.0001** |
| Ringing in the ears | 55 (47.8) | 103 (71.5) | **0.0001** | 184 (46.0) | 228 (58.3) | **0.0006** |
| Shortness of breath | 49 (42.6) | 76 (52.8) | 0.11 | 139 (34.8) | 222 (56.9) | **<0.0001** |
| Dry eyes | 36 (31.3) | 82 (56.9) | **<0.0001** | 130 (32.5) | 221 (56.5) | **<0.0001** |
| Depressed mood | 69 (60.0) | 87 (60.4) | 0.99 | 220 (55.0) | 212 (54.4) | 0.89 |
| Nervousness | 53 (46.1) | 81 (56.3) | 0.13 | 185 (46.3) | 212 (54.4) | **0.023** |
| Blurred vision | 43 (37.4) | 82 (56.9) | **0.002** | 137 (34.3) | 208 (53.2) | **<0.0001** |
| Heartburn | 35 (30.4) | 70 (48.6) | **0.003** | 118 (29.5) | 199 (51.0) | **<0.0001** |
| Frequent urination | 32 (27.8) | 81 (56.3) | **<0.0001** | 134 (33.5) | 194 (49.7) | **<0.0001** |
| Sun sensitivity | 24 (20.9) | 66 (45.8) | **<0.0001** | 97 (24.3) | 193 (49.4) | **<0.0001** |
| Increased sweating | 31 (27.0) | 73 (50.7) | **0.0001** | 153 (38.3) | 191 (48.8) | **0.003** |
| Dry mouth | 30 (26.1) | 81 (56.3) | **<0.0001** | 115 (28.8) | 190 (48.6) | **<0.0001** |
| Frequent loose stools | 41 (35.7) | 74 (51.4) | **0.012** | 115 (28.8) | 180 (46.2) | **<0.0001** |
| Hair loss | 28 (24.3) | 82 (56.9) | **<0.0001** | 100 (25.0) | 180 (46.0) | **<0.0001** |
| Loss of appetite | 43 (37.4) | 81 (56.3) | **0.003** | 145 (36.3) | 175 (44.9) | **0.003** |
| Decreased sex drive | 44 (38.3) | 63 (43.8) | 0.38 | 129 (32.3) | 173 (44.2) | **0.0006** |
| Hives | 16 (13.9) | 70 (48.6) | **<0.0001** | 38 (9.5) | 168 (43.0) | **<0.0001** |
| Rash | 20 (17.4) | 61 (42.4) | **<0.0001** | 42 (10.5) | 147 (37.6) | **<0.0001** |
| Hearing difficulties | 19 (16.5) | 50 (34.7) | **0.001** | 87 (21.8) | 126 (32.2) | **0.001** |
| Bladder cramps | 18 (15.7) | 52 (36.1) | **0.0002** | 65 (16.3) | 119 (30.5) | **<0.0001** |
| Oral ulcers | 17 (14.8) | 41 (28.5) | **0.011** | 57 (14.3) | 119 (30.4) | **<0.0001** |
| Wheezing | 3 (2.6) | 36 (25.0) | **<0.0001** | 20 (5.0) | 98 (25.1) | **<0.0001** |
| Loss of/change in taste | 6 (5.2) | 31 (21.5) | **0.0001** | 21 (5.3) | 63 (16.1) | **<0.0001** |
| None | 3 (2.6) | 1 (0.7) | 0.33 | 27 (6.8) | 5 (1.3) | **<0.0001** |

<sup>a</sup> All data shown as $n$ (%). <sup>b</sup> $P$ values result from Fisher's test for categorical data and Kruskal-Wallis rank sum test for numeric data. <sup>c</sup> Order of conditions based on highest to lowest percentage in HSD Hi MC group. <sup>d</sup> Bold indicates significant value. <sup>e</sup> Abbreviations: TMJ, temporomandibular joint.

We found that more hEDS/HSD patients with a Hi MC score had a diagnosis of fibromyalgia based on the questionnaire vs. those with a lo MC score (hEDS OR 3.70, CI 2.06-6.89, *p* < 0.0001; HSD OR 3.38, CI 2.41-4.73, *p* < 0.0001) (**Table 9**, **Supplementary Table 9**). Similarly, more patients with a Hi MC score reported fibromyalgia symptoms/comorbidities than those with a Lo MC score except for ‘depressed mood’ (hEDS *p* = 0.99, HSD *p* = 0.89), which would not be expected to be affected by MC activity. All other fibromyalgia-related symptoms were increased in patients with a Hi MC score in either hEDS or HSD. The top 5 fibromyalgia-related symptoms/comorbidities that were significantly increased in patients with Hi vs. Lo MC scores for both diagnoses included numbness (hEDS OR 2.98, CI 1.73-5.11, *p* < 0.0001; HSD OR 2.61, CI 1.89-3.56, *p* < 0.0001), easy bruising (hEDS OR 4.19, CI 2.29-7.59, *p* < 0.0001; HSD OR 2.69, CI 1.98-3.64, *p* < 0.0001), nausea (hEDS OR 2.42, CI 1.38-4.20, *p* = 0.002; HSD OR 2.47, CI 1.82-3.35, *p* < 0.0001), multiple sensitivities (hEDS OR 5.42, CI 3.17-9.38, *p* < 0.0001; HSD OR 4.23, CI 3.15-5.68, *p* < 0.0001) and heat intolerance (hEDS OR 3.16, CI .91-5.19, *p* < 0.0001; HSD OR 3.21, CI 2.38-4.29, *p* < 0.0001) (**Table 9**, **Supplementary Table 9**). Thus, more hEDS/HSD patients with a high MC score report fibromyalgia symptoms and comorbidities.

### 3.10 Psychological conditions and abuse

When we examined psychological symptoms and comorbidities, we found that only a few psychological conditions were associated with a high MC score. We found that hEDS/HSD patients with a Hi vs. Lo MC score reported more anxiety (hEDS OR 2.25, CI 1.33-3.92, *p* = 0.004; HSD OR 1.41, CI 1.02-1.93, *p* = 0.041), suggesting that MC activity may make anxiety worse (**Table 10**, **Supplemental Table 10**). However, we did not find that MC activity based on the MC score affected depression (hEDS *p* = 0.24, HSD *p* = 0.99). We also found that more patients with hEDS and a Hi MC score reported obsessive-compulsive disorder (OCD) whereas patients with HSD did not report this association (hEDS OR 2.87, CI 1.45-5.55, *p* = 0.001; HSD OR 1.05, CI 0.73-1.51, *p* = 0.85).

**Table 10.** Psychological conditions in patients diagnosed with hEDS or HSD with low (Lo) vs. high (Hi) MC scores (*n* = 1,050)

| Condition | hEDS<sup>a</sup><br>Lo MC<br>($n = 115$) | hEDS<br>Hi MC<br>($n = 144$) | *P* value<sup>b</sup> | HSD<br>Lo MC<br>($n = 400$) | HSD<sup>c</sup><br>Hi MC<br>($n = 391$) | *P* value<sup>b</sup> |
| --- | --- | --- | --- | --- | --- | --- |
| Anxiety | 70 (60.9) | 112 (77.8) | **0.004<sup>d</sup>** | 285 (71.3) | 304 (77.7) | **0.041** |
| Depression | 68 (59.1) | 96 (66.7) | 0.24 | 249 (62.3) | 244 (62.4) | 0.99 |
| Abuse | 35 (30.4) | 84 (58.3) | **<0.0001** | 134 (33.5) | 202 (51.7) | **<0.0001** |
| PTSD<sup>e</sup> | 21 (18.3) | 71 (49.3) | **<0.0001** | 89 (22.3) | 150 (38.4) | **<0.0001** |
| OCD | 14 (12.2) | 41 (28.5) | **0.001** | 66 (16.5) | 67 (17.1) | 0.85 |
| Panic disorder | 11 (9.6) | 27 (18.8) | 0.05 | 51 (12.8) | 67 (17.1) | 0.09 |
| Eating disorders | 13 (11.3) | 24 (16.7) | 0.28 | 60 (15.0) | 63 (16.1) | 0.69 |
| Body dysmorphic disorder | 6 (5.2) | 13 (9.0) | 0.34 | 32 (8.0) | 42 (10.7) | 0.22 |
| Bipolar disorder | 5 (4.3) | 6 (4.2) | 0.99 | 26 (6.5) | 22 (5.6) | 0.66 |
| Personality disorder | 5 (4.3) | 4 (2.8) | 0.52 | 13 (3.3) | 11 (2.8) | 0.84 |
| Conversion disorder | 0 (0.0) | 4 (2.8) | 0.13 | 4 (1.0) | 8 (2.0) | 0.26 |
| Schizophrenia | 0 (0.0) | 0 (0.0) | - | 0 (0.0) | 1 (0.3) | 0.49 |
| Other | 7 (6.1) | 10 (6.9) | 0.99 | 18 (4.5) | 38 (9.7) | **0.005** |
| Unknown | 7 (6.1) | 3 (2.1) | 0.11 | 13 (3.3) | 7 (1.8) | 0.26 |
| No issues | 27 (23.5) | 18 (12.5) | **0.031** | 69 (17.3) | 52 (13.3) | 0.14 |
| **Type of abuse** |  |  |  |  |  |  |
| Emotional/verbal abuse | 23 (20.0) | 72 (50.0) | **<0.0001** | 109 (27.3) | 165 (42.2) | **<0.0001** |
| Physical abuse | 14 (12.2) | 53 (36.8) | **<0.0001** | 60 (15.0) | 123 (31.5) | **<0.0001** |
| Sexual abuse | 20 (17.4) | 54 (37.5) | **0.0005** | 71 (17.8) | 115 (29.4) | **0.0001** |
| Unknown | 4 (3.5) | 4 (2.8) | 0.99 | 9 (2.3) | 18 (4.6) | 0.08 |
| **ACE score** | $n = 71$ | $n = 91$ | **0.008** | $n = 132$ | $n = 146$ | **0.009** |
| mean (SD) | 2.48<br>(2.49) | 3.59<br>(2.86) |  | 2.57<br>(2.51) | 3.32<br>(2.54) |  |

<sup>a</sup> All data shown as $n$ (%). <sup>b</sup> *P* values result from Fisher's test for categorical data and Kruskal-Wallis rank sum test for numeric data. <sup>c</sup> Order of conditions based on highest to lowest percentage in HSD Hi MC group. <sup>d</sup> Bold indicates significant value. <sup>e</sup> Abbreviations: OCD, obsessive-compulsive disorder; PTSD, post-traumatic stress disorder.

In contrast, we found a large increase in the percentage of patients with hEDS/HSD that reported abuse and post-traumatic stress disorder (PTSD) if they had a Hi vs. Lo MC score-with values almost doubling (Abuse: hEDS Lo MC 30.4% vs. Hi MC 58.3%, OR 3.20, CI 1.93-5.31, *p* < 0.0001; HSD Lo MC 33.5% vs. Hi MC 51.7%, OR 2.12, CI 1.59-2.81, *p* < 0.0001; PTSD: hEDS Lo MC 18.3% vs. Hi MC 49.3%, OR 4.35, CI 2.48-7.64, *p* < 0.0001; HSD Lo MC 22.3% vs. Hi MC 38.4%, OR 2.18, CI 1.59-2.97, *p* < 0.0001) (**Table 10, Supplementary Table 10, Fig3a**). Individual types of abuse for both diagnoses were also almost 2x higher in patients with Hi vs. Lo MC scores (emotional/verbal abuse: hEDS Lo MC 20.0% vs. Hi MC 50.0%, OR 4.00, CI 2.25-7.15, *p* < 0.0001; HSD Lo MC 27.3% vs. Hi MC 42.2%, OR 1.95, CI 1.45-2.61, *p* < 0.0001; physical abuse: hEDS Lo MC 12.2% vs. Hi MC 36.8%, OR 4.20, CI 2.19-7.96, *p* < 0.0001; HSD Lo MC 15.0% vs. Hi MC 31.5%, OR 2.60, CI 1.83-3.67, *p* < 0.0001; sexual abuse: hEDS Lo MC 17.4% vs. Hi MC 37.5%, OR 2.85, CI 1.58-5.16, *p* = 0.0005; HSD Lo MC 17.8% vs. Hi MC 29.4%, OR 1.93, CI 1.39-2.69, *p* < 0.0001) (**Table 10, Supplementary Table 10, Fig3a**). ORs with CIs for high vs. low MC scores for abuse, types of abuse and PTSD are shown in Figure 3a.

**Figure 3.**
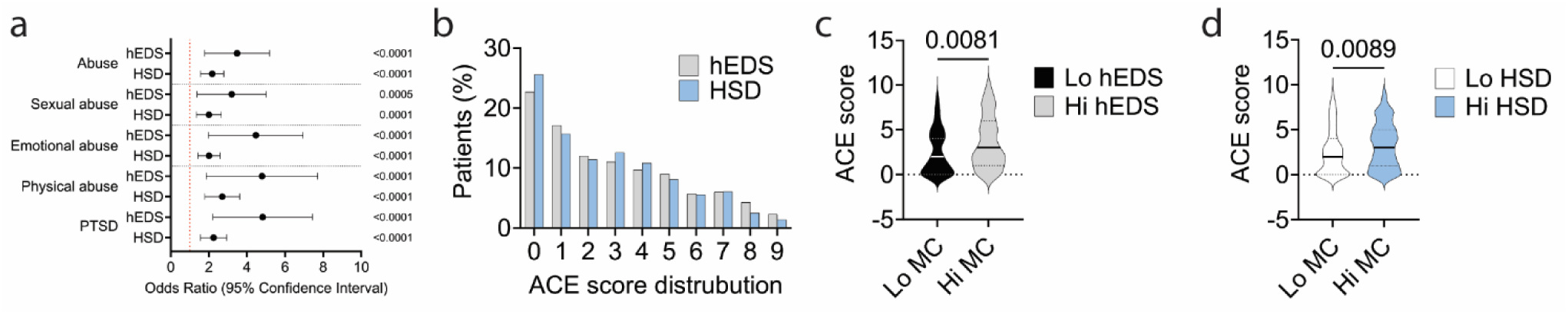
Patients with high MC scores self-report more abuse, PTSD and ACE scores. a) Odds ratios (OR) and 95% confidence interval (CI) comparing low vs. high MC scores in patients with hEDS and HSD. P values obtained from Fisher’s exact test. b) Distribution of the adverse childhood events (ACE) score for patients with hEDS or HSD. ACE score in c) hEDS and d) HSD patients with low (Lo) vs. high (Hi) MC scores. P values derived from Student’s *t* test for parametric data and Mann-Whitney rank test for nonparametric data.

Because we observed that hEDS/HSD patients with a high MC score reported more abuse and PTSD, we examined adverse childhood experiences (ACE) questions from our Intake Questionnaire. Figure 3b shows the distribution of ACE scores for hEDS and HSD. We found that hEDS/HSD patients with a Hi vs. Lo MC scores had higher ACE scores (hEDS: Lo MC 2.48 ±2.49 vs. Hi MC 3.59 ±2.86, OR 2.61, CI 1.23-5.44, *p* = 0.008; HSD: Lo MC 2.57 ±2.51 vs. Hi MC 3.32 ±2.54, OR 1.98, CI 1.09-3.54, *p* = 0.009) (**Table 10, Supplementary Table 10, Fig3c,d**).

## 4. Discussion

There is general agreement among hEDS and HSD practitioners that patients have evidence of widespread MC activity based on many MC-associated symptoms/comorbidities. However, the criteria (consensus-1) for a diagnosis of MCAS and/or a MCAD is quite stringent,^14^ essentially requiring that an anaphylactic response be captured clinically which is frequently difficult to obtain. In contrast, the consensus-2 criteria for MCAS is mainly based on a list of widespread symptoms/comorbidities making it difficult to define or measure.^17^ For this reason, we (Fairweather, Bruno) devised a novel MC score based on self-reported MC-mediated responses including environmental allergies, asthma (past and/or present), atopy, food allergies, venom/stinging insect allergies, rhinitis, eczema, drug/chemical allergies, MCAS, MC hyperactivity, and/or a tryptase mutation which could serve in addition to the MCAS criteria to quantify widespread MC issues. Based on the population distribution of the MC score we separated patients into those with a low MC score (0-1) vs. those with a high MC score (≥5). Although the MC score is self-reported and may not accurately reflect clinical diagnoses of these 11 conditions, the purpose of the score was to quantify widespread MC-related symptoms with the goal of providing a simple, fast and low-cost method for determining the severity of MC-related symptoms in patients with hEDS, HSD or other conditions. The MC score is intended to be used in addition to MCAS criteria. The small 11-question questionnaire could be sent through the patient portal, filled out in the waiting room, or administered during consultation.

Importantly, we found that hEDS/HSD patients with high vs. low MC scores reported widespread MC-related symptoms/comorbidities in many body regions. We found that 80.7% of hEDS and 95.6% of HSD patients reported 135 symptoms/comorbidities (typically found in these patients^25–28^) more often if they had a high MC score. Interestingly, the symptoms/comorbidities reported to be associated with MCAS by expert consensus by Akin et al. and Valent et al. were all reported more often in hEDS/HSD patients with high MC scores,^13,14^ indicating overlap between MCAS and a high MC score for these symptoms. We were able to validate the MC score in part by demonstrating higher serum total IgE levels by ELISA in a subset of hEDS/HSD patients indicating MC-specific IgE-mediated responses. However, it is highly likely that the widespread symptoms associated with a high MC score are not restricted to IgE-mediated pathology but include the myriad of other MC produced mediators which we plan to investigate in the future.

In contrast, a high MC score was not reflected in higher clinical random blood tryptase or urinary MC mediators analyzed from the medical record in a subset of patients. Interestingly, we found that a high MC score significantly reduced urinary creatinine levels in hEDS patients, which are used to normalize the data, indicating that those values were not accurate. Creatinine levels can be affected if patients have diseases that affect kidneys like diabetes, chronic kidney disease, or systemic lupus erythematosus and should be taken into consideration. From the medical record analysis, we had two hEDS patients with high MC scores with conditions that may have affected their kidney health including one with kidney stones and another with May Thurner Syndrome, but no other apparent kidney issues. Another possibility is that patients with hEDS/HSD report frequent urinary infections which we found were increased in hEDS/HSD patients with high MC scores (**Table 7**).

All the blood and urinary samples we examined in this study were collected on the day of the patient’s appointment at the EDS Clinic and so were ‘random’ rather than targeting an acute or anaphylactic MC event. Although theoretically possible, it is practically quite difficult for patients to provide a sample in 1-4 hours after they experience an acute event, even with a physician’s order. Thus, it is difficult for patients at our Clinic to obtain a diagnosis of MCAS. We found that around 50% of hEDS/HSD patients with clinical tryptase levels above the background level received a diagnosis of MCAS in our subset analysis. It is also possible that MC issues are so widespread and frequent in hEDS/HSD patients that the MCs are ‘exhausted’ creating a high ‘background’ value and difficulty in obtaining a ‘high’ anaphylactic value for MC markers like tryptase. For unclear reasons, we also infrequently find that urinary MC markers are elevated during our routine clinical assessment. Yet the results of this study indicate that around 25% of the patients seen at the EDS Clinic have widespread MC issues based on having a high MC score. This percentage is similar to the percentage that has been suggested for the prevalence of MCAS using consensus-2.^17^ Our findings demonstrate the utility of a MC score to identify patients with widespread MC symptoms/comorbidities and quantify the severity level of their MC activity.

We previously reported that MC symptoms are often self-reported in patients with hEDS/HSD.^27,28^ Aside from several meeting abstracts, there are few studies examining the relationship of MC activation to hEDS/HSD in the literature aside from Brock et al.^10^ Our finding of overlap in allergy/mast cell symptoms in hEDS and HSD is consistent with the report by Ritelli et al. who described plasma biomarkers in both hEDS and HSD, but not in controls, that are the product of ECM degradation (i.e., collagen I and fibronectin fragments) which is a primary role of MCs.^6^ A recent study by Griggs et al. is also consistent with our findings.^35^ Grigg et al. conducted proteomics on the blood of 29 female hEDS patients vs. 29 controls and found that hEDS patients had evidence of immune dysregulation particularly in complement pathway proteins.^35^ Complement and MC activation are closely intertwined with end-products of the alternative complement cascade iC3b, C3a and C5a activating MCs via CD11b (complement receptor/CR3) and anaphylaxis receptors C3aR and C5aR.^36,37^ A large survey study by Daly et al. of hEDS (*n* = 3,360) and HSD (*n* = 546) patients, with diagnoses verified by survey questions but not physicians, reported widespread symptoms and comorbidities for most body systems compared to All of Us data.^38^ They found that 70-80% of hEDS/HSD patients self-reported allergies.^38^ For several MC-related symptoms like MCAS, chronic sinusitis, and chronic urticaria they found that hEDS patients reported more MC-related symptoms than patients with HSD.^38^ In this study, we did not directly compare hEDS to HSD patients. Overall, we found very similar responses between hEDS and HSD patients for MC-related symptoms and comorbidities in the subgroup of patients with widespread MC issues based on a high vs. low MC score. Thus, our paper is the first to create a MC score to assess/quantify widespread MC issues in patients with hEDS/HSD in a large number of patients with clinician-verified diagnoses of hEDS and HSD using the 2017 diagnostic criteria.^4,29^

Another interesting finding of this study was which self-reported symptoms/conditions did not appear to have a relationship with a high MC score. These included POTS (patients self-reported this diagnosis), depression/depressed mood, ulcerative colitis and Crohn’s disease, and several neurological tests/conditions including autism spectrum disorder (ASD), intracranial hypertension, and cerebral spinal fluid (CSF) leak. Aside from depression, other psychological conditions that were not reported more often in hEDS/HSD patients with a high MC score included eating disorders, body dysmorphic disorder, bipolar disorder, personality disorder, conversion disorder and schizophrenia.

In this study, around 60-80% of hEDS/HSD patients self-reported that they had been diagnosed with POTS, but the percentage was not increased in patients with high MC activation suggesting that MC activation may not be associated with POTS. However, these percentages likely reflect patient perceptions, and future studies need to be conducted in hEDS/HSD patients with physician-confirmed diagnoses. Interestingly, the percentage of patients who said they had been assessed for POTS was around 30-40%. Although several review articles and systematic reviews examine whether a relationship exists between MC activation and POTS in hEDS/HSD patients,^22,39^ little data exists. Wang et al. examined 195 patients seen for autonomic dysfunction and found that MCAS occurred in 31% of EDS patients with POTS vs. 2% in the non-POTS/EDS group.^11^ In contrast, our study suggests that no relationship exists between patients with widespread MC activation (high MC score) and POTS, but more research is needed. Depression is often reported in patients with hEDS/HSD. In this study, around 60-67% of hEDS/HSD patients self-reported depression/depressed mood, which did not increase in patients with a high MC score suggesting that MC activation is not affecting depression. A recent review in *Nature Reviews Disease Primers* on major depressive disorder indicates that inflammation via the stress pathway has a role in the pathogenesis of disease but does not specifically mention MCs.^40^ However, several studies suggest a link between high MC activation and the development of depression.^41,42^ More research is needed on this topic.

However, psychologically associated conditions that were reported more often in hEDS/HSD patients with high MC activation included anxiety, PTSD and abuse-emotional/verbal, sexual and physical. We did not obtain information on the perceived source of the PTSD, but many hEDS/HSD patients describe medically associated PTSD related to difficulty obtaining a diagnosis and/or dismissal of their symptoms by many physicians.^43^ A meta-analysis from 24 countries reported a pooled national prevalence estimate of child sexual abuse of 15% of girls and 8% of boys.^44^ In the US, at least 1 in 4 girls (25%) and 1 in 20 boys (5%) experience childhood sexual abuse.^44^ We found that 30% of hEDS/HSD patients with low MC scores reported sexual abuse compared to around 50% with high MC scores. A relationship between stress and MC activation is well established in the literature. MCs have been shown to express corticotropin-releasing hormone (CRH) receptors, which bind CRH that is released by the hypothalamic-pituitary-adrenal (HPA) axis in response to stress.^45–47^ Additionally, a higher ACE score has been associated with the development of inflammatory conditions later in life.^24,33^ Future research should examine whether hEDS/HSD patients benefit from targeted anxiety/abuse/PTSD psychotherapy and MC inhibitors.

There are several limitations to this study. Symptoms/comorbidities including the MC score were self-reported and not validated through another method such as checking the medical record for diagnostic codes. Future studies are needed to examine whether the key findings of this study can be verified using larger cohorts. The site of the study is a tertiary care center and findings from this study may not be generalizable to other regions of the US or world. A strength of this study is that hEDS/HSD patients were diagnosed with hEDS or HSD using the most recent 2017 criterion by physician experts in a large cohort of patients. An additional strength of the study is the development of a MC score to quantitate widespread MC issues and to correlate to ACE scores. It is important to note that the MC score is intended to be used in addition to MCAS criteria-not to replace it. Of note, the diagnostic criteria for hEDS and HSD are currently under revision. Until revised diagnostic criteria for hEDS and HSD are published, hEDS and HSD remain distinct diagnoses and so are examined in this study according to the 2017 diagnostic criteria.

In conclusion, this is the first study to develop a method for quantifying widespread MC symptoms. We show that hEDS/HSD patients with a high MC score not only self-report more MC-specific symptoms/comorbidities but also widespread symptoms throughout the body demonstrating its usefulness as a clinical and research tool to better understand MC symptoms in hEDS and HSD patients.

## 6 Author Contributions

DF designed the study. DF and KAB developed the MC score. RMB, AMZ, DRTK, SNG diagnosed hEDS and HSD patients. FCW, AAD, KAB and DF built and/or managed the electronic database of patient data. FCW, JJF, AAD, CH, MWS, AMP, CJH, KAB and DF collected data. FCW, DJZ, LER, JJF, AAD, SNWR, KAB, DF analyzed the data. FCW, DJZ, LER, JJF, AAD, KAS, SMM, DVD, AGE, SCS, IS, BWEW, PSA, CLS, RMB, AMZ, DRTK, SNG, KAB and DF interpreted the data. DF wrote the manuscript. FCW and DJZ assisted with writing the paper. DRTK, SNG, KAB and DF agree to be accountable to all aspects of the work ensuring that questions related to accuracy and integrity of any part of the work are properly investigated and resolved. All authors edited and revised the paper for important content. All authors approved the final version of the paper.

## Data Availability

All data produced in the present study are available upon reasonable request to the authors.

## 8 Acknowledgments

None.

## 10 Supplementary Material

Supplementary material is associated with this article.

## 11 Data Availability Statement

The original contributions presented in the study are included in the article; further inquiries can be directed to the corresponding author.

