## Supplemental data for "Mast cell score associates with wide-spread mast cell symptoms and comorbidities in patients with hEDS and HSD"

### Supplementary Material

### Tables

Supplemental Table 1. Environmental exposure demographics of patients diagnosed with hEDS or HSD with low (Lo) vs. high (Hi) MC scores (*n* = 1,050)

| Condition | hEDS[a] Lo MC | hEDS Hi MC | *P* value[b] | HSD Lo MC | HSD Hi MC | *P* value[b] |
|---|---|---|---|---|---|---|
| ***BMI*** | | | **0.024** | | | **<0.0001** |
| *n* | 104 | 130 | | 370 | 365 | |
| Median (interquartile range) | 22.4 (20.3-26.1) | 24.6 (21.3-29.1) | | 24.4 (21.3-29.7) | 27.5 (23.3-33.0) | |
| ***Smoking history (n)*** | 115 | 144 | | 400 | 390 | |
| Yes-Currently | 16 (13.9) | 11 (7.6) | 0.11 | 37 (9.3) | 33 (8.5) | 0.71 |
| Yes-Past | 14 (12.2) | 27 (18.8) | 0.17 | 43 (10.8) | 70 (17.9) | **0.004** |
| No | 85 (73.9) | 106 (73.6) | 0.99 | 313 (78.3) | 286 (73.3) | 0.11 |
| Unknown | 0 (0.0) | 0 (0.0) | 0.99 | 7 (1.8) | 1 (0.3) | 0.07 |
| ***Number of cigarettes smoked/day (n)*** | 40 | 36 | | 80 | 103 | |
| 1-5 | 4 (10.0) | 7 (19.4) | 0.33 | 15 (18.8) | 22 (21.4) | 0.71 |
| 6-10 | 2 (5.0) | 6 (16.7) | 0.13 | 3 (3.8) | 13 (12.6) | **0.038** |
| 10-20 | 3 (7.5) | 9 (25.0) | 0.06 | 5 (6.3) | 15 (14.6) | 0.09 |
| >20 | 2 (5.0) | 0 (0.0) | 0.49 | 2 (2.5) | 7 (6.8) | 0.30 |
| Unknown | 11 (27.5) | 3 (8.3) | **0.040** | 11 (13.8) | 13 (12.6) | 0.83 |
| Vaping | 18 (45.0) | 11 (30.6) | 0.24 | 44 (55.0) | 33 (32.0) | **0.003** |
| ***Secondary smoke exposure(n)*** | 115 | 144 | | 400 | 390 | |
| Yes | 49 (42.6) | 58 (40.3) | 0.79 | 138 (34.5) | 184 (47.2) | **0.0003** |
| No | 64 (55.7) | 85 (59.0) | 0.61 | 256 (64.0) | 201 (51.5) | **0.0004** |
| Unknown | 2 (1.7) | 1 (0.7) | 0.59 | 6 (1.5) | 5 (1.3) | 0.99 |
| ***Alcohol consumption (n)*** | 115 | 144 | | 400 | 390 | |
| Yes-Currently | 59 (51.3) | 82 (56.9) | 0.38 | 194 (48.5) | 190 (48.7) | 0.99 |
| Yes-Past | 22 (19.1) | 41 (28.5) | 0.11 | 89 (22.3) | 122 (31.3) | **0.005** |
| No | 33 (28.7) | 21 (14.6) | **0.009** | 115 (28.8) | 76 (19.5) | **0.003** |
| Unknown | 1 (0.9) | 0 (0.0) | 0.44 | 2 (0.5) | 2 (0.5) | 0.99 |
| ***Number of alcoholic drinks consumed on average/week (n)*** | 75 | 123 | | 274 | 306 | |
| 0-1 | 29 (38.7) | 56 (45.5) | 0.38 | 126 (46.0) | 123 (40.2) | 0.18 |
| 2-3 | 14 (18.7) | 18 (14.6) | 0.55 | 41 (15.0) | 54 (17.6) | 0.43 |
| 4-7 | 5 (6.7) | 6 (4.9) | 0.75 | 26 (9.5) | 19 (6.2) | 0.16 |
| Rarely | 24 (32.0) | 39 (31.7) | 0.99 | 76 (27.7) | 100 (32.7) | 0.21 |
| Unknown | 3 (4.0) | 4 (3.3) | 0.99 | 5 (1.8) | 10 (3.3) | 0.31 |
| ***Alcohol exposure before birth (n)*** | 115 | 144 | | 400 | 391 | |
| Yes | 3 (2.6) | 6 (4.2) | 0.74 | 9 (2.3) | 13 (3.3) | 0.39 |
| No | 106 (92.2) | 124 (86.1) | 0.16 | 350 (87.5) | 321 (82.1) | **0.037** |


This is a provisional file, not the final typeset article

| | | | | | | |
|---|---|---|---|---|---|---|
| Unknown | 6 (5.2) | 14 (9.7) | 0.24 | 41 (10.3) | 57 (14.6) | 0.07 |
| ***Illicit drug use (n)*** | 115 | 133 | | 400 | 390 | |
| Yes-Currently | 3 (2.6) | 2 (1.5) | 0.67 | 23 (5.8) | 13 (3.3) | 0.12 |
| Yes-Past | 2 (1.7) | 14 (10.5) | **0.008** | 19 (4.8) | 16 (4.1) | 0.73 |
| No | 103 (89.6) | 117 (88.0) | 0.84 | 337 (84.3) | 338 (86.7) | 0.36 |
| Unknown/ choose not to disclose | 7 (6.1) | 11 (8.3) | 0.63 | 21 (5.3) | 23 (5.9) | 0.76 |
| ***Drug Exposure as a baby*** | 115 | 144 | | 400 | 391 | |
| Yes | 2 (1.7) | 6 (4.2) | 0.31 | 10 (2.5) | 11 (2.8) | 0.83 |
| No | 104 (90.4) | 117 (81.3) | 0.05 | 347 (86.8) | 323 (82.6) | 0.11 |
| Unknown | 9 (7.8) | 21 (14.6) | 0.12 | 43 (10.8) | 57 (14.6) | 0.11 |

[a] Most data shown as *n* (%). [b] P values obtained from Fisher's test for categorical data and Mann-Whitney test for numeric data. [c] Bold indicates significant value.

Supplemental Table 2. Odds ratios (OR) and 95% confidence intervals (CI) for MC-related symptoms that comprise the MC score (*n* = 1,050)

| **Lo vs. Hi MC** | **hEDS OR (95% CI)** | ***P* value[a]** | **HSD [b] OR (95% CI)** | ***P* value[a]** |
|---|---|---|---|---|
| Environmental allergies | 439.3 (107.1-1859) | **<0.0001**[c] | 128.3 (68.86-244.9) | **<0.0001** |
| Atopy | 536.3 (63.30-5466) | **<0.0001** | 108.0 (28.69-318.8) | **<0.0001** |
| Drug allergies | 46.34 (21.93-92.94) | **<0.0001** | 29.05 (19.77-42.92) | **<0.0001** |
| Rhinitis (i.e., hayfever) | 87.24 (33.59-208.5) | **<0.0001** | 170.3 (91.03-318.6) | **<0.0001** |
| Food allergies | 38.92 (14.98-92.20) | **<0.0001** | 55.34 (33.44-93.71) | **<0.0001** |
| Asthma | 29.28 (11.98-67.03) | **<0.0001** | 56.19 (30.78-105.8) | **<0.0001** |
| Eczema | 21.60 (9.28-51.17) | **<0.0001** | 14.54 (9.64-21.74) | **<0.0001** |
| Venom allergies | 24.46 (7.64-76.18) | **<0.0001** | 44.56 (18.79-102.5) | **<0.0001** |
| MCAS | 20.47 (6.85-63.92) | **<0.0001** | 45.71 (15.61-133.9) | **<0.0001** |
| Overactive mast cells | ∞ (4.15-∞) | **<0.0001** | 42.09 (8.26-214.6) | **<0.0001** |
| Tryptase mutation | ∞ (0.37-∞) | 0.50 | 13.51 (0.76-240.6) | **0.014** |

[a] P values obtained from Fisher's exact test. [b] Order of conditions based on highest to lowest percentage in HSD Hi MC group from Table 2. [c] Bold indicates significant value $p<0.05$; values higher than $p<0.0004$ were not significant after Bonferroni correction.

Supplementary Table 3. Odds ratios (OR) and 95% confidence intervals (CI) for MC-related symptoms listed in Akin et al.[13] and Valent et al.[14] ($n$ = 1,050)

| **Hi MC v. Lo MC** | **hEDS OR (95% CI)** | ***P* value[a]** | **HSD[b] OR (95% CI)** | ***P* value[a]** |
|---|---|---|---|---|
| Headache | 5.74 (3.74-8.72) | **0.0001**[c] | 4.63 (3.09-6.99) | **<0.0001** |
| Nasal congestion | 42.20 (18.31-90.32) | **<0.0001** | 12.41 (8.33-18.14) | **<0.0001** |
| Dizzy | 1.73 (0.92-3.30) | 0.10 | 2.74 (1.92-3.92) | **<0.0001** |
| Migraine | 3.72 (2.14-6.44) | **<0.0001** | 4.14 (2.93-5.81) | **<0.0001** |
| Nausea | 2.42 (1.38-4.20) | **0.0005** | 2.47 (1.82-3.35) | **<0.0001** |
| Pruritus (itching) | 7.46 (4.10-13.17) | **<0.0001** | 7.69 (5.40-11.02) | **<0.0001** |
| Abdominal cramping | 2.01 (1.20-3.39) | **0.0093** | 2.27 (1.69-3.05) | **<0.0001** |
| Flushing | 8.97 (4.83-16.34) | **<0.0001** | 4.48 (3.21-6.26) | **<0.0001** |
| Tachycardia | 2.64 (1.51-4.58) | **0.0004** | 2.85 (2.06-3.94) | **<0.0001** |
| Urticaria (hives) | 10.69 (5.67-20.54) | **<0.0001** | 9.96 (6.70-15.04) | **<0.0001** |
| Diarrhea | 3.19 (1.82-5.47) | **<0.0001** | 3.32 (2.39-4.63) | **<0.0001** |
| Difficulty breathing | 3.75 (2.13-6.50) | **<0.0001** | 4.66 (3.18-6.89) | **<0.0001** |
| Throat swelling | 13.28 (6.40-26.51) | **<0.0001** | 5.74 (3.74-8.72) | **<0.0001** |
| Hypotension | 3.03 (1.69-5.27) | **0.0002** | 3.08 (2.10-4.49) | **<0.0001** |
| Vomiting | 1.94 (1.08-3.55) | **0.040** | 3.30 (2.19-4.98) | **<0.0001** |
| Wheezing | 12.44 (4.04-39.25) | **<0.0001** | 6.38 (3.89-10.54) | **<0.0001** |
| Weal/blisters | 8.07 (2.78-21.75) | **<0.0001** | 5.90 (3.37-10.48) | **<0.0001** |
| Collapse | 1.92 (1.04-3.47) | **0.049** | 1.66 (1.10-2.51) | **0.018** |

[a] P values obtained from Fisher's exact test. [b] Order of conditions based on highest to lowest percentage in HSD Hi MC group from Table 3. [c] Bold indicates significant value $p<0.05$; values higher than $p<0.0004$ were not significant after Bonferroni correction.

Supplementary Table 4. Odds ratios (OR) and 95% confidence intervals (CI) for skin symptoms and comorbidities ($n$ = 1,050)

| **Hi MC v. Lo MC** | **hEDS OR (95% CI)** | ***P* value[a]** | **HSD[b] OR (95% CI)** | ***P* value[a]** |
|---|---|---|---|---|
| Easy bruising | 3.16 (1.39-7.51) | **0.006**[c] | 2.01 (1.36-2.99) | **0.0005** |
| Easy scarring | 2.05 (1.05-3.97) | **0.045** | 2.05 (1.49-2.81) | **<0.0001** |
| Strech marks | 1.94 (1.12-3.30) | **0.025** | 1.96 (1.46-2.62) | **<0.0001** |
| Poor wound healing | 3.01 (1.73-5.25) | **<0.0001** | 2.26 (1.69-3.01) | **<0.0001** |
| Abnormal scarring | 2.50 (1.49-4.15) | **0.0004** | 3.03 (2.26-4.05) | **<0.0001** |
| Eczema | 20.30 (8.63-45.61) | **<0.0001** | 10.95 (7.25-16.48) | **<0.0001** |
| Rosacea | 0.72 (0.35-1.61) | 0.43 | 2.59 (1.73-3.89) | **<0.0001** |
| Psoriasis | 2.01 (0.90-4.74) | 0.12 | 4.42 (2.47-8.05) | **<0.0001** |

[a] P values obtained from Fisher's exact test. [b] Order of conditions based on highest to lowest percentage in HSD Hi MC group from Table 3. [c] Bold indicates significant value $p<0.05$; values higher than $p<0.0004$ were not significant after Bonferroni correction.



This is a provisional file, not the final typeset article

Supplementary Table 5. Odds ratios (OR) and 95% confidence intervals (CI) for joint issues ($n$ = 1,050)

| **Hi MC v. Lo MC** | **hEDS OR (95% CI)** | ***P* value**[a] | **HSD**[b] **OR (95% CI)** | ***P* value**[a] |
|---|---|---|---|---|
| Joint pain | 4.27 (1.69-10.72) | **0.002**[c] | 4.02 (1.91-8.53) | **0.0002** |
| Joint subluxations | 4.43 (2.15-8.82) | **<0.0001** | 2.33 (1.63-3.30) | **<0.0001** |
| Joint sprains | 4.38 (2.36-7.93) | **<0.0001** | 3.77 (2.72-5.22) | **<0.0001** |
| Joint dislocations | 2.38 (1.44-3.95) | **0.001** | 1.87 (1.37-2.54) | **<0.0001** |

[a] P values obtained from Fisher's exact test. [b] Order of conditions based on highest to lowest percentage in HSD Hi MC group from Table 3. [c] Bold indicates significant value $p<0.05$; values higher than $p<0.0004$ were not significant after Bonferroni correction.

Supplementary Table 6. Odds ratios (OR) and 95% confidence intervals (CI) for GI issues ($n$ = 1,050)

| **Hi MC v. Lo MC** | **hEDS OR (95% CI)** | ***P* value**[a] | **HSD**[b] **OR (95% CI)** | ***P* value**[a] |
|---|---|---|---|---|
| Nausea | 2.57 (1.51-4.35) | **0.0005**[c] | 2.51 (1.85-3.41) | **<0.0001** |
| Constipation | 2.86 (1.67-4.79) | **<0.0001** | 2.44 (1.83-3.27) | **<0.0001** |
| Diarrhea | 2.40 (1.46-4.02) | **0.0009** | 2.78 (2.08-3.70) | **<0.0001** |
| GERD[d] | 3.75 (2.21-6.45) | **<0.0001** | 2.95 (2.20-3.96) | **<0.0001** |
| IBS | 2.86 (1.67-4.79) | **0.0001** | 2.83 (2.11-3.81) | **0.0001** |
| Dyspepsia | 3.39 (1.90-6.10) | **<0.0001** | 2.71 (2.00-3.68) | **<0.0001** |
| Vomiting | 2.26 (1.35-3.80) | **0.003** | 2.41 (1.78-3.27) | **<0.0001** |
| Hemorrhoids | 2.94 (1.72-4.92) | **<0.0001** | 2.31 (1.68-3.17) | **<0.0001** |
| Gastroparesis | 3.45 (1.73-6.98) | **0.0002** | 1.89 (1.33-2.66) | **0.0004** |
| Hernias | 2.22 (1.24-4.06) | **0.0091** | 2.62 (1.81-3.79) | **<0.0001** |
| Anal fissure | 2.41 (1.25-4.71) | **0.011** | 3.13 (1.97-4.98) | **<0.0001** |
| Fecal incontinence | 4.34 (1.29-14.38) | **0.014** | 1.83 (0.98-3.41) | 0.06 |
| Rectal prolapse | 1.93 (0.78-4.97) | 0.19 | 2.26 (1.10-4.62) | **0.024** |
| Barrett's esophagus | 4.10 (0.55-48.71) | 0.23 | 4.39 (1.10-17.55) | **0.020** |
| Ulcerative colitis | 1.44 (0.44-4.42) | 0.76 | 2.52 (0.72-8.81) | 0.14 |
| Crohn's disease | 1.61 (0.18-23.45) | 0.99 | 2.69 (0.62-11.63) | 0.17 |
| Other | 3.19 (1.60-6.22) | **0.0009** | 1.99 (1.34-3.00) | **0.0009** |

[a] P values obtained from Fisher's exact test. [b] Order of conditions based on highest to lowest percentage in HSD Hi MC group from Table 3. [c] Bold indicates significant value $p<0.05$; values higher than $p<0.0004$ were not significant after Bonferroni correction. [d] Abbreviations: GERD, gastroesophageal reflux disease; IBS, irritable bowel syndrome.

Supplementary Table 7. Odds ratios (OR) and 95% confidence intervals (CI) for genitourinary issues ($n$ = 1,050)

| **Hi MC v. Lo MC** | **hEDS OR (95% CI)** | ***P* value**[a] | **HSD**[b] **OR (95% CI)** | ***P* value**[a] |
|---|---|---|---|---|
| Dyspareunia | 3.39 (1.92-5.96) | **<0.0001**[c] | 2.63 (1.90-3.64) | **<0.0001** |
| Recurrent urinary tract infection | 2.56 (1.49-4.52) | **0.001** | 2.06 (1.50-2.84) | **<0.0001** |
| Incontinence | 2.45 (1.32-4.58) | **0.005** | 2.72 (1.93-3.79) | **<0.0001** |
| Pelvic floor dysfunction | 3.84 (1.99-7.29) | **<0.0001** | 3.45 (2.32-5.21) | **<0.0001** |
| PCOS | 3.50 (1.64-7.77) | **0.001** | 2.23 (1.54-3.26) | **<0.0001** |
| Recurrent yeast infection | 2.15 (1.06-4.34) | **0.031** | 2.52 (1.71-3.69) | **<0.0001** |
| Endometriosis | 3.03 (1.59-5.59) | **0.0006** | 2.74 (1.86-4.03) | **<0.0001** |
| Pelvic floor spasm | 2.18 (1.05-4.75) | 0.05 | 4.37 (2.61-7.39) | **<0.0001** |
| Interstitial cystitis | 2.49 (1.00-6.55) | **0.043** | 5.41 (2.88-10.62) | **<0.0001** |
| Recurrent vaginal bacteria infection | 1.22 (0.55-2.79) | 0.68 | 1.93 (1.17-3.21) | **0.012** |
| Uterine prolapse | 2.18 (0.79-5.68) | 0.22 | 2.33 (1.09-4.76) | **0.030** |
| Bladder prolapse | 2.369 (0.88-6.11) | 0.15 | 2.92 (1.35-6.29) | **0.009** |

[a] P values obtained from Fisher's exact test. [b] Order of conditions based on highest to lowest percentage in HSD Hi MC group from Table 3. [c] Bold indicates significant value $p<0.05$; values higher than $p<0.0004$ were not significant after Bonferroni correction. [d] Abbreviations: PCOS, polycystic ovary syndrome.



This is a provisional file, not the final typeset article

Supplementary Table 8. Odds ratios (OR) and 95% confidence intervals (CI) for neurological conditions ($n$ = 1,050)

| **Hi MC v. Lo MC** | **hEDS OR (95% CI)** | ***P* value[a]** | **HSD[b] OR (95% CI)** | ***P* value[a]** |
|---|---|---|---|---|
| Brain fog | 3.56 (2.25-5.53) | **0.0003**[c] | 2.15 (1.51-3.08) | **<0.0001** |
| Headache | 1.39 (0.73-2.64) | 0.33 | 2.05 (1.46-2.84) | **<0.0001** |
| Migraine | 2.94 (1.78-4.82) | **<0.0001** | 3.57 (2.65-4.82) | **<0.0001** |
| Tinnitus | 3.67 (2.21-6.06) | **<0.0001** | 1.50 (1.13-1.98) | **0.006** |
| Vertigo | 2.32 (1.41-3.80) | **0.002** | 2.35 (1.74-3.16) | **<0.0001** |
| Autonomic dysfunction | 2.56 (1.53-4.26) | **0.0004** | 2.71 (2.00-3.67) | **<0.0001** |
| Chronic migraine | 3.20 (1.79-5.77) | **<0.0001** | 2.45 (1.79-3.35) | **<0.0001** |
| Assessed POTS[d] | 1.83 (1.09-3.06) | **0.027** | 2.01 (1.48-2.73) | **<0.0001** |
| ADD/ADHD | 1.26 (0.72-2.16) | 0.42 | 1.49 (1.11-2.00) | **0.009** |
| Neuropathy | 4.00 (2.14-7.39) | **<0.0001** | 2.83 (2.03-3.91) | **<0.0001** |
| NDPH | 1.23 (0.73-2.03) | 0.51 | 2.06 (1.53-2.79) | **<0.0001** |
| Cluster headache | 2.24 (1.12-4.54) | **0.022** | 2.17 (1.50-3.13) | **<0.0001** |
| Current or past abnormal brain MRI | 1.73 (0.87-3.45) | 0.16 | 1.66 (1.08-2.58) | **0.023** |
| ASD | 1.80 (0.83-4.01) | 0.14 | 1.39 (0.90-2.18) | 0.15 |
| Intracranial hypertension | 1.21 (0.31-3.86) | 0.99 | 1.88 (0.97-3.64) | 0.07 |
| Arnold Chiari malformation | 2.03 (0.43-10.35) | 0.47 | 2.00 (1.02-3.92) | **0.046** |
| CSF leak | 1.62 (0.422-6.02) | 0.74 | 1.66 (0.74-3.57) | 0.24 |

[a] P values obtained from Fisher's exact test. [b] Order of conditions based on highest to lowest percentage in HSD Hi MC group from Table 3. [c] Bold indicates significant value $p<0.05$; values higher than $p<0.0004$ were not significant after Bonferroni correction. [d] Abbreviations: ADD, attention deficit disorder; ADHD, attention deficit hyperactivity disorder; ASD, autism spectrum disorder; CSF, cerebral spinal fluid; MRI, magnetic resonance imaging; NDPH, new daily persistent headache; POTS, postural orthostatic tachycardia syndrome.

Supplementary Table 9. Odds ratios (OR) and 95% confidence intervals (CI) for fibromyalgia conditions ($n$ = 1,050)

| **Hi MC v. Lo MC** | **hEDS OR (95% CI)** | ***P* value**[a] | **HSD**[b] **OR (95% CI)** | ***P* value**[a] |
|---|---|---|---|---|
| Fibromyalgia diagnosis | 3.70 (2.06-6.89) | **<0.0001**[c] | 3.38 (2.41-4.73) | **<0.0001** |
| Headache | 1.57 (0.78-3.05) | 0.22 | 2.26 (1.56-3.26) | **<0.0001** |
| Numbness | 2.98 (1.73-5.11) | **<0.0001** | 2.61 (1.89-3.56) | **<0.0001** |
| Easy bruising | 4.19 (2.29-7.59) | **<0.0001** | 2.69 (1.98-3.64) | **<0.0001** |
| Lightheadedness | 1.59 (0.90-2.86) | 0.14 | 1.91 (1.40-2.59) | **<0.0001** |
| Nausea | 2.42 (1.38-4.20) | **0.002** | 2.47 (1.82-3.35) | **<0.0001** |
| Multiple sensitivities | 5.42 (3.17-9.38) | **<0.0001** | 4.23 (3.15-5.68) | **<0.0001** |
| Heat intolerance | 3.16 (1.91-5.19) | **<0.0001** | 3.21 (2.38-4.29) | **<0.0001** |
| Pain lower abdomen | 2.01 (1.20-3.34) | **0.009** | 2.27 (1.69-3.05) | **<0.0001** |
| Sense of imbalance | 1.71 (1.04-2.83) | **0.041** | 2.52 (1.87-3.38) | **<0.0001** |
| Muscle weakness | 1.83 (1.11-3.03) | **0.021** | 2.21 (1.65-2.96) | **<0.0001** |
| Constipation | 2.19 (1.31-3.63) | **0.002** | 2.32 (1.73-3.10) | **<0.0001** |
| Bowel cramps | 2.02 (1.21-3.31) | **0.006** | 3.12 (2.33-4.18) | **<0.0001** |
| Palpitations | 3.26 (1.96-5.39) | **<0.0001** | 1.97 (1.49-2.63) | **<0.0001** |
| TMJ disorder[d] | 3.27 (1.92-5.34) | **<0.0001** | 2.25 (1.69-2.97) | **<0.0001** |
| Cold intolerance | 2.23 (1.34-3.77) | **0.003** | 2.39 (1.80-3.16) | **<0.0001** |
| Chest discomfort | 2.15 (1.29-3.56) | **0.003** | 2.07 (1.56-2.75) | **<0.0001** |
| Ringing in the ears | 2.74 (1.65-4.49) | **0.0001** | 1.64 (1.23-2.17) | **0.0006** |
| Shortness of breath | 1.51 (0.91-2.44) | 0.11 | 2.48 (1.86-3.29) | **<0.0001** |
| Dry eyes | 2.90 (1.76-4.78) | **<0.0001** | 2.70 (2.02-3.60) | **<0.0001** |
| Depressed mood | 1.02 (0.62-1.66) | 0.99 | 0.97 (0.74-1.28) | 0.89 |
| Nervousness | 1.50 (0.91-2.43) | 0.13 | 1.38 (1.04-1.83) | **0.023** |
| Blurred vision | 2.22 (1.33-3.67) | **0.002** | 2.18 (1.64-2.89) | **<0.0001** |
| Heartburn | 2.16 (1.31-3.56) | **0.003** | 2.49 (1.85-3.34) | **<0.0001** |
| Frequent urination | 3.34 (1.99-5.63) | **<0.0001** | 1.97 (1.47-2.61) | **<0.0001** |
| Sun sensitivity | 3.21 (1.81-5.64) | **<0.0001** | 3.05 (2.25-4.13) | **<0.0001** |
| Increased sweating | 2.79 (1.66-4.73) | **0.0001** | 1.54 (1.17-2.05) | **0.003** |
| Dry mouth | 3.64 (2.15-6.26) | **<0.0001** | 2.34 (1.74-3.15) | **<0.0001** |
| Frequent loose stools | 1.91 (1.14-3.16) | **0.012** | 2.12 (1.59-2.86) | **<0.0001** |
| Hair loss | 4.11 (2.39-6.89) | **<0.0001** | 2.56 (1.89-3.46) | **<0.0001** |
| Loss of appetite | 2.15 (1.29-3.56) | **0.003** | 2.15 (1.29-3.56) | **0.003** |
| Decreased sex drive | 1.26 (0.77-2.06) | 0.38 | 1.67 (1.25-2.22) | **0.0006** |
| Hives | 5.85 (3.11-10.99) | **<0.0001** | 7.18 (4.89-10.59) | **<0.0001** |
| Rash | 3.49 (1.95-6.28) | **<0.0001** | 5.14 (3.49-7.51) | **<0.0001** |
| Hearing difficulties | 2.69 (1.46-5.01) | **0.001** | 1.71 (1.25-2.36) | **0.001** |
| Bladder cramps | 3.05 (1.64-5.44) | **0.0002** | 2.26 (1.61-3.20) | **<0.0001** |
| Oral ulcers | 2.29 (1.23-4.30) | **0.011** | 2.63 (1.86-3.77) | **<0.0001** |
| Wheezing | 12.44 (4.04-39.25) | **<0.0001** | 6.38 (3.89-10.54) | **<0.0001** |
| Loss of/change in taste | 4.98 (2.09-11.72) | **0.0001** | 3.47 (2.09-5.77) | **<0.0001** |

[a] P values obtained from Fisher's exact test. [b] Order of conditions based on highest to lowest percentage in HSD Hi MC group from Table 3. [c] Bold indicates significant value $p<0.05$; values higher than $p<0.0004$ were not significant after Bonferroni correction. [d] Abbreviations: TMJ, temporomandibular joint.



This is a provisional file, not the final typeset article

Supplementary Table 10. Odds ratios (OR) and 95% confidence intervals (CI) for psychological conditions ($n$ = 1,050)

| **Hi MC v. Lo MC** | **hEDS OR (95% CI)** | ***P* value**[a] | **HSD**[b] **OR (95% CI)** | ***P* value**[a] |
|---|---|---|---|---|
| Anxiety | 2.25 (1.33-3.92) | **0.004**[c] | 1.41 (1.02-1.93) | **0.041** |
| Depression | 1.38 (0.83-2.29) | 0.24 | 1.01 (0.75-1.35) | 0.99 |
| Abuse | 3.20 (1.93-5.31) | **<0.0001** | 2.12 (1.59-2.81) | **<0.0001** |
| PTSD[d] | 4.35 (2.48-7.64) | **<0.0001** | 2.18 (1.59-2.97) | **<0.0001** |
| OCD | 2.87 (1.45-5.55) | **0.001** | 1.05 (0.73-1.51) | 0.85 |
| Panic disorder | 2.18 (1.05-4.75) | 0.05 | 1.42 (0.95-2.10) | 0.09 |
| Eating disorders | 1.57 (0.77-3.12) | 0.28 | 1.09 (0.74-1.61) | 0.69 |
| Body dysmorphic disorder | 1.80 (0.70-4.79) | 0.34 | 1.38 (0.86-2.22) | 0.22 |
| Bipolar disorder | 0.96 (0.27-2.83) | 0.99 | 0.86 (0.49-1.54) | 0.66 |
| Personality disorder | 0.63 (0.19-2.15) | 0.52 | 0.86 (0.39-1.91) | 0.84 |
| Conversion disorder | ∞ (0.79-∞) | 0.13 | 1.95 (0.62-6.18) | 0.26 |
| Schizophrenia | - | - | 3.08 (0.13-75.76) | 0.49 |
| Other | 1.15 (0.41-2.98) | 0.99 | 2.29 (1.29-4.13) | 0.005 |
| Unknown | 0.33 (0.09-1.22) | 0.11 | 0.56 (0.23-1.38) | 0.26 |
| No issues | 0.47 (0.24-0.89) | **0.031** | 0.74 (0.49-1.09) | 0.14 |
| **Type of abuse** | | | 0.00 (0.00-0.00) | |
| Emotional/verbal abuse | 4.00 (2.25-7.15) | **<0.0001** | 1.95 (1.45-2.61) | **<0.0001** |
| Physical abuse | 4.20 (2.19-7.96) | **<0.0001** | 2.60 (1.83-3.67) | **<0.0001** |
| Sexual abuse | 2.85 (1.58-5.16) | **0.0005** | 1.93 (1.39-2.69) | **0.0001** |
| Unknown | 0.79 (0.23-2.78) | 0.99 | 2.04 (0.92-4.52) | 0.08 |
| **ACE score** | 2.61 (1.23-5.44) | **0.008** | 1.98 (1.09-3.54) | **0.009** |

[a] P values obtained from Fisher's exact test. [b] Order of conditions based on highest to lowest percentage in HSD Hi MC group from Table 3. [c] Bold indicates significant value $p<0.05$; values higher than $p<0.0004$ were not significant after Bonferroni correction. [d] Abbreviations: OCD, obsessive-compulsive disorder; PTSD, post-traumatic stress disorder.
